# Prevalence, determinants, and impact on general health and working capacity of post-acute sequelae of COVID-19 six to 12 months after infection: a population-based retrospective cohort study from southern Germany

**DOI:** 10.1101/2022.03.14.22272316

**Authors:** Raphael S. Peter, Alexandra Nieters, Hans-Georg Kräusslich, Stefan O. Brockmann, Siri Göpel, Gerhard Kindle, Uta Merle, Jürgen M. Steinacker, Dietrich Rothenbacher, Winfried V. Kern, the EPILOC Phase 1 Study Group

**Author notes:** **Author for correspondence** Prof. Dr. Winfried V. Kern, Division of Infectious Diseases, University Medical Centre, Hugstetter Straße 55 · D-79106 Freiburg, Germany. Co-first authors (Raphael S. Peter, Alexandra Nieters). Joint senior authors (Dietrich Rothenbacher, Winfried V. Kern). **Contributors** WVK led the study conceptualisation and the development of the research question, supported by HGK, AN, RP. WVK, AN and DR supervised the study. SB and GK contributed to the design of the study. RP, AN and DR were involved in data acquisition and statistical analysis. SB, SG, UM and JS contributed to the acquisition and interpretation of the data. WVK, AN, RP and DR had full access to and verified the data, take responsibility for data integrity and the accuracy of the data analysis, and for the decision to submit for publication. All authors were involved in drafting or critically revising the manuscript, and all authors approved the final version. **Declaration of interests** We declare no competing interests. **Data sharing** Data from EPILOC Phase 1 are publicly available for research purposes upon request. **Funding** This work was funded by the Baden-Württemberg Federal State Ministry of Science and Art (grant number MR/S028188/1) and the German pension fund (“Deutsche Rentenversicherung”) Baden-Württemberg.

## Abstract

**Background:** Post-acute sequelae of SARS-CoV-2 infection have commonly been described after COVID-19, but few population-based studies have examined symptoms six to 12 months after acute SARS-CoV-2 infection and their associations with general health recovery and working capacity.

**Methods:** This population-based retrospective cohort study in four geographically defined regions in southern Germany included persons aged 18-65 years with PCR confirmed SARS-CoV-2 infection between October 2020 and March 2021. Symptom frequencies (six to 12 months after versus before acute infection, expressed as prevalence differences [PD] and prevalence ratios [PR]), symptom severity and clustering, risk factors and associations with general health recovery, and working capacity were analysed.

**Findings:** Among a total of 11 710 subjects (mean age 44·1 years, 59·8% females, 3·5% previously admitted with COVID-19, mean follow-up time 8.5 months) the most prevalent symptoms with PDs >20% and PRs >5% were rapid physical exhaustion, shortness of breath, concentration difficulties, chronic fatigue, memory disturbance, and altered sense of smell. Female sex and severity of the initial infection were the main risk factors. Prevalence rates, however, appeared substantial among both men and women who had a mild course of acute infection, and PCS considerably affected also younger subjects. Fatigue (PD 37·2%) and neurocognitive impairment (PD 31·3%) as symptom clusters contributed most to reduced health recovery and working capacity, but chest symptoms, anxiety/depression, headache/dizziness and pain syndromes were also prevalent and relevant for working capacity, with some differences according to sex and age. When considering new symptoms with at least moderate impairment of daily life and ≤80% recovered general health or working capacity, the overall estimate for post-COVID syndrome was 28·5% (age- and sex-standardised rate 26·5%).

**Interpretation:** The burden of self-reported post-acute symptoms and possible sequelae, notably fatigue and neurocognitive impairment, remains considerable six to 12 months after acute infection even among young and middle-aged adults after mild acute SARS-CoV-2 infection, and impacts general health and working capacity.

**Research in context:** *Evidence before this study:* Previous studies have shown that post-acute sequelae of COVID-19 are common, in particular among patients who had been admitted to hospital for COVID-19. Post-acute self-reported complaints and symptoms often are diverse, nonspecific and sometimes of unknown severity and functional relevance. We searched PubMed and medRxiv for studies published between January 2021 and February 2022, using search terms describing “long covid, post-acute sequelae of COVID-19, prevalence, and systematic review”, with no language restrictions. Searches with the terms “long covid”, “post-acute sequelae of COVID-19”, “post-covid condition” and “post-covid syndrome” were also done in PROSPERO, and we screened the website of the UK Office for National Statistics (www.ons.gov.uk) for long covid studies. We found more than 20 systematic reviews summarising post-acute symptom patterns among adults and a prevalence of “any” or “defined” symptoms (such as respiratory symptoms or symptoms related to mental health) or of medically assessed functional impairment (pulmonary or cardiac or neurocognitive function). Two reviews reported of health-related quality of life assessments. The prevalence of post-acute sequelae of COVID-19 or long covid/post-covid syndromes ranged between <10 to >70%, in part due to lack of uniform and clear case definitions, variable follow-up times, and non-inclusion of outpatients with initially mild disease. Most papers reviewed presented high heterogeneity and had a short follow-up, and there were very few papers estimating the prevalence of post-covid syndrome beyond six months after acute infection. The studies with the largest number of subjects were either including only patients after hospital admission, used online surveys of subjects with self-reported suspected and confirmed COVID-19 or electronic medical records only. We found one (small but) comprehensive population-based study from Switzerland assessing post-covid syndrome prevalence and associations with quality of life and health recovery with a follow-up time ranging from six to 10 months. Two further population-based studies from Switzerland and Norway investigated long covid symptoms among subgroups with ≥6 months (n=498) and 11 to 12 months (n=170) of follow-up after acute infection, respectively.

*Added value of this study:* With this large population-based study, we provide evidence of persistence of new symptom clusters (not present before acute infection) such as fatigue, neurocognitive impairment, chest symptoms, smell or taste disorder, and anxiety/depression beyond six months after acute infection, with a prevalence of >20% for each of these five clusters. We show that the three most frequent clusters (fatigue, neurocognitive impairment, chest symptoms) are often interfering with daily life and activities, often co-occur, and that both fatigue and neurocognitive impairment have the largest impact on working capacity, while long-term smell and taste disorders are reported relatively independent of other complaints. Age in this 18-65-year old adult population was not a major determinant of symptom prevalence, but we confirm severity of the initial infection and female sex as consistent risk factors for various manifestations of medium-term post-COVID syndrome, and age as risk factor for self-reported reduced working capacity, which overall and at population level exceeded 10%.

*Implications of all the available evidence:* Future research should include the medical validation of the key symptom clusters of post-COVID syndrome, determine the possible causes, and urgently address prognostic factors and therapeutic options. The described key symptom clusters contributed most to reduced general health status and working capacity in middle-aged adults. The findings of this study may also help develop a more consistent and relevant definition of post-COVID syndrome with major implications for research and medical practice.

## Introduction

Severe acute respiratory syndrome coronavirus 2 (SARS-CoV-2) has caused the COVID-19 viral pandemic with far-reaching consequences, including a worldwide health crisis. Although respiratory infection is the primary clinical manifestation, COVID-19 is considered a multi-organ systemic disease that includes the lung, heart, vascular system, brain and other organ systems.^1,2^ Most infections are mild or even asymptomatic, especially among children and adolescents, while the likelihood for severe disease and the need for hospital admission increase substantially with age and comorbidity.^3,4^ The 30-day mortality among hospitalised cases from Germany in a nationwide claims data cohort study (first wave) was 24% overall and 53% among patients requiring ventilation.^5^

Besides the acute phase morbidity and mortality, post-acute health problems and sequelae have been reported in COVID-19 survivors. According to a review, up to 80% of COVID-19 patients continue to complain about health problems following acute infection, and more than 50 adverse effects were reported.^6^ The pathophysiology of many post-acute symptoms has remained unresolved. Symptoms can last for weeks and represent delayed reconvalescence or can persist or recur even three months or longer into the post-acute phase.^6-9^ While “long covid” has been defined as ongoing symptoms beyond four weeks after acute infection, post-COVID-19 condition or post-COVID syndrome (PCS) is considered in individuals with symptoms lasting for at least two months, being unexplained by an alternative diagnosis, and occurring three months from the acute infection.^10^ So far, very few larger-scaled studies have examined the symptomatology and prevalence of PCS beyond six months after acute infection and its association with health-related quality of life, well-being, and working capacity in a population-based, non-clinical sample.

The primary aims of the present study were to describe symptoms and symptom clusters of PCS six to 12 months after acute infection, estimate their prevalence, describe risk factors and examine the association of symptom clusters with general health and working capacity. The data were generated in a large population-based study in southern Germany involving 18 to 65-year-old subjects with PCR-confirmed SARS-CoV-2 infection notified to local health authorities.

## Material and methods

EPILOC (*<u>Epi</u>*demiology of *<u>Lo</u>*ng *<u>C</u>*ovid) is a non-interventional retrospective cohort study conducted in four administratively and geographically defined regions in the Federal State of Baden-Württemberg in southwestern Germany. The study included subjects aged 18-65 years who were tested positive in a SARS-CoV-2 PCR between October 1^st^, 2020 and April 1^st^, 2021 and whose infection was notified (according to the German Infection Protection Act) to the local public health authorities responsible for the following four regions: Freiburg (city of Freiburg, district of Breisgau-Hochschwarzwald, district of Emmendingen), Heidelberg (city of Heidelberg, Rhein-Neckar district), Tübingen (city of Tübingen, city of Reutlingen, Zollernalb district), and Ulm (city of Ulm, Alb-Donau district, district of Heidenheim, district of Biberach) – regions with a total population of 2·7 million combined.

Surviving persons were directly contacted by the local public health authorities via postal mail between late August and September 2021. All study materials (i.e. participant information, informed consent form, and a standardised questionnaire) were included in the letter. Subjects were asked to provide written informed consent and send the study materials (postage-paid) to the trustee office of the study centre at the Freiburg University Medical Centre. The trustee separated the declaration of informed consent from the completed questionnaire and forwarded the questionnaires to the data management centre at Ulm University.

The study was conducted according to the Declaration of Helsinki, and ethical approval was obtained from the respective ethical review boards of the study centres in Freiburg (21/1484) and Ulm (337/21). It is registered with DRKS (“Deutsches Register Klinischer Studien”), (DRKS 00027012), and the present analysis follows the STROBE recommendations.

### Data sources and measurements

The standardised questionnaire included sociodemographic characteristics, lifestyle factors, and medically attended comorbidities already present before the acute SARS-CoV-2 infection. It questioned the presence of thirty specific symptoms before and during the acute infection phase as well as at the time of filling out the questionnaire (i.e. six to 12 months after acute infection) by yes/no responses. Further new or ongoing current symptoms could be added in a free-text field. If any of the symptoms were present at the time of the survey, we asked for associated medical treatment (yes/no) and whether and to which grade each symptom impaired daily life and activities (“how much do you feel impaired by this at the moment?”) using a 4-point Likert scale (none, light, moderate, or strong).

For the evaluation of fatigue (already included in the list of symptoms), we additionally used the 10-item Fatigue Assessment Scale (FAS).^11^ A threshold score of ≥22 is used for determining the presence of substantial fatigue, and a threshold score of ≥35 for extreme fatigue. To assess working capacity we adapted questions from the short form of the work ability index.^12^ Participants assessed their current general health recovery and current working capacity compared with the situation before the acute SARS-CoV-2 infection on a 10-point Likert scale (10% steps from 0% to 100%). To evaluate the current health-related quality of life (HRQoL), we used the SF-12 (short form-12) questionnaire assessing physical and mental health-related quality of life components (https://www.rand.org/health-care/surveys_tools/mos/12-item-short-form.html).

### Statistical methods

The characteristics of the study population were evaluated descriptively. We obtained the relative frequency of the individual symptoms prior to, during acute infection, and at the time of the survey (i.e. six to 12 months after index infection) and calculated the differences in prevalence current versus before the acute infection (PD) and the relative prevalence ratios (PR) (current versus before acute infection), including a 95% confidence interval (CI). Gender- and age-stratified representations were also provided.

Clusters of strongly correlated current symptoms (not present prior to the SARS-CoV-2 infection) were identified using exploratory polychoric factor analysis. The identified symptom clusters were visualised by means of a co-occurrence network using Gephi 0.9.2. We repeated the analysis based on symptoms of grade moderate or strong as sensitivity analysis.

The PRs for symptom clusters (with 95% CIs) by possible determinants (age, gender, education, smoking status, BMI, time since positive PCR, severity of acute infection, and pre-existing conditions) were calculated mutually adjusted. The association of each current symptom cluster with loss of general health and working capacity compared to pre-infection was computed (adjusted for the presence of other symptom clusters) and expressed as attributable loss (in percentages). Corresponding 95% CIs for the attributable impairment/loss were estimated using a parametric bootstrap.

Prevalence, PRs, and PDs were estimated using Poisson models. All CIs are based on robust standard errors, accounting for possible dispersion and the correlated nature of the data in case of comparing symptoms before and after acute infection. Statistical procedures were performed with the SAS statistical software package (release 9.4 SAS Institute Inc.) or R version 4.1.2.

## Results

A total of 50,457 adults with confirmed SARS-CoV-2 infection were invited to participate in the study, of whom 12,053 (24%) subjects responded, and 11,710 provided at least information on age and sex (see study flow-chart in the **appendix, figure S1)**. The mean time between the initial positive PCR test and the time of the survey was 8·5 months.

As shown in **table 1**, the mean age of the participants was 44·1 years (SD 13·7), and there were slightly more females (59·8%) than males. The majority of subjects were born in Germany (88·8%), had German nationality (94·2%), were from urban areas (84·3%), and had a university entrance qualification (51·9%). More than half of the participants reported pre-pandemic full-time employment (56·8%). Reported chronic pre-existing health conditions included musculoskeletal disorders (32·5%), cardiovascular disorders (20·5%), neurological and sensory disorders (18·7%), and respiratory diseases (15·2%) besides others. The vast majority of subjects (77·5%) did not require medical care for the previous acute SARS-CoV-2 infection, 19·0% reported outpatient care, and less than 4% had required hospital admission **(table 1)**.

**Table 1.** Characteristics of the study population.

|  | N | Number (%) or<br>mean (standard<br>deviation) |
| --- | --- | --- |
| Age (years), mean (sd) | 11 710 | 44.1 (13.7) |
| Age class (years), N (%) |  |  |
| <30 |  | 2474 (21.1) |
| 30 - <40 | 11 710 | 2158 (18.4) |
| 40 - <50 |  | 2075 (17.7) |
| 50 - <60 |  | 3443 (29.4) |
| ≥ 60 |  | 1560 (13.3) |
| Sex, N (%) |  |  |
| Male | 11710 | 4829 (41.2) |
| Female |  | 6881 (58.8) |
| Marital status, N (%) |  |  |
| Single |  | 3425 (29.8) |
| Married/living together | 11 492 | 7563 (65.8) |
| Living apart |  | 368 (3.2) |
| Widowed |  | 136 (1.2) |
| University entrance qualification, N (%) |  |  |
| Yes | 11 678 | 6065 (51.9) |
| No |  | 5613 (48.1) |
| Place of birth, N (%) |  |  |
| Germany | 11 668 | 10 355 (88.8) |
| Other |  | 1313 (11.3) |
| Nationality, N (%) |  |  |
| German | 11 688 | 11 004 (94.2) |
| Other* |  | 684 (5.9) |
| Place of residence, N (%) |  |  |
| Mostly urban | 11 365 | 7246 (63.8) |
| Partly urban |  | 2329 (20.5) |
| Mostly rural |  | 1790 (15.8) |
| Pre-pandemic employment, N (%) |  |  |
| Full time |  | 6608 (56.8) |
| Part time | 11 628 | 3220 (27.7) |
| Studying/vocational education |  | 1143 (9.8) |
| None |  | 657 (5.7) |
| Current employment, N (%) |  |  |
| Full time |  | 6335 (54.4) |
| Part time | 11 651 | 3215 (27.6) |
| Studying/vocational education |  | 1031 (8.9) |
| None |  | 1070 (9.2) |
| Smoking status, N (%) |  |  |
| Current | 11 678 | 1192 (10.2) |
| Former |  | 2882 (24.7) |
| Never |  | 7604 (65.1) |
| BMI (kg/m<sup>2</sup>), mean (sd) | 11 619 | 26.1 (5.3) |
| Obese (≥30 kg/m<sup>2</sup>), N (%) | 11 619 | 2171 (18.7) |
| Pre-existing conditions, N (%) |  |  |
| Musculoskeletal disorders (including rheumatism) | 11 448 | 3717 (32·5) |
| Cardiovascular disorders (including hypertension) | 11 477 | 2347 (20·5) |
| Neurological or sensory disorders | 11 480 | 2146 (18·7) |
| Metabolic disorders | 11 554 | 2139 (18·5) |
| Mental disorders | 11 479 | 1911 (16·7) |
| Respiratory diseases | 11 467 | 1747 (15·2) |
| Dermatological diseases | 11 547 | 1511 (13·1) |
| Cancer | 11 323 | 433 ( 3·8) |
| Time since positive PCR test (months), mean (sd) | 11 521 | 8·5 (1·6) |
| Treatment of acute SARS-CoV-2 infection, N (%) |  |  |
| No medical care |  | 8988 (77·5) |
| Outpatient care | 11 602 | 2202 (19·0) |
| Inpatient care (without ICU) |  | 315 (2·7) |
| Intensive care |  | 97 (0·8) |

### Prevalence of individual symptoms

The reported symptom prevalence at the three time points, including PRs and PDs for all queried symptoms, are depicted in **figure 1**. Six to 12 months after acute infection, concentration difficulties, memory disturbance, altered sense of smell, shortness of breath, chronic fatigue, and rapid physical exhaustion were the most frequent symptoms in relative and absolute measures compared to pre-infection, with PRs >5 and PDs >20%, respectively. Other post-acute individual symptoms – all with PDs between 10 and 20% – were dizziness, altered sense of taste, and sleep disorder. With PDs below 3%, vomiting, nausea, abdominal pain, diarrhoea, chills, fever, skin rash, and others contributed little to the PCS symptomatology as did free-text added post-acute symptoms (most common: abnormal heart beat and disturbed vision) that were mentioned by <1% of the respondents (free-text data not shown).

**Figure 1.**
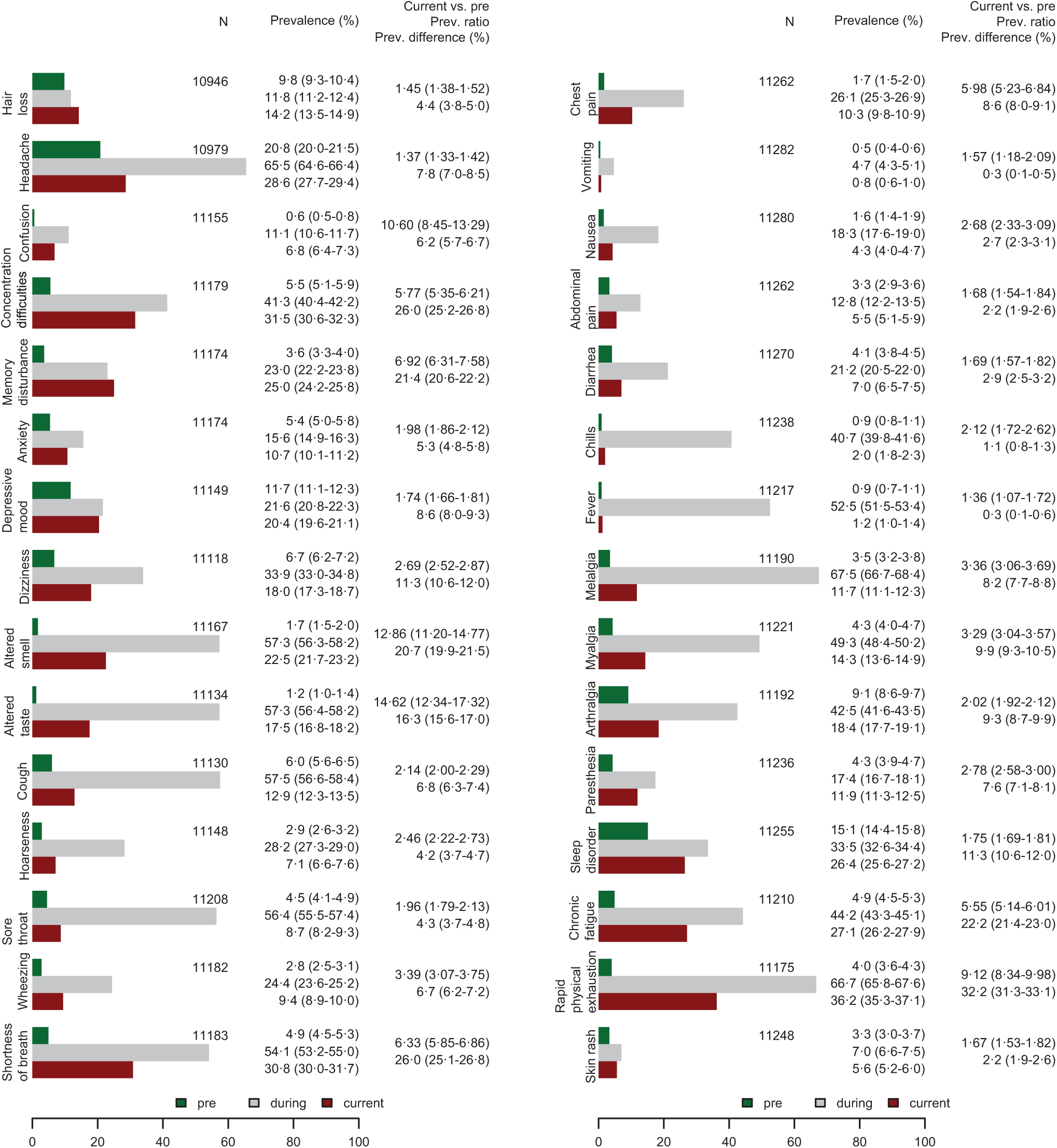
Prevalence of symptoms in percent (%) before (pre), during, and 6-12 months after (current) the SARS-CoV-2 index infection including prevalence ratio (current divided by pre) and prevalence difference (current minus pre) with 95 % confidence intervals. N represents the number of respondents in the respective age strata.

Further prevalence measures according to age categories and stratified to sex are shown in the **appendix, figure S2**. Interestingly, symptoms prior to the acute infection and – more so – post-acute were reported more frequently by women, with PDs for hair loss, headache and nausea differing by a factor greater than two between sexes. In addition, subjects in the age groups 40 to 59 years were most affected by many symptoms, and we observed notable increases in PDs for many symptoms with age, while the prevalences and PDs for altered sense of smell decreased with age.

We noted differences between current symptoms regarding the grade of impairment, showing higher grades of impairment for women for most symptoms (**see appendix, figure S3**). Among the participants reporting any symptom at six to 12 months, 11·2% reported some medical treatment for their symptoms, most often (in absolute numbers) for shortness of breath, rapid physical exhaustion and chronic fatigue – with minor differences between men and women (**see appendix, figure S3**). The proportion of participants with at least one symptom of any grade of impairment was 63·7%, compared with a rate of 41·5% for symptoms of moderate or strong grade.

### Symptom clusters

Several of the 30 post-acute symptoms were strongly correlated and could be combined into 13 symptom clusters **(figure 2)**. The individual symptoms rapid physical exhaustion and chronic fatigue, for example, were combined into the cluster “fatigue” which was the most common symptom cluster (37·2%), followed by “neurocognitive impairment” with a prevalence of 31·3%, “chest symptoms” (30·2%), “smell or taste disorder” (23·6%), and “anxiety/depression” (21·1%). This ranking remained similar when only symptoms with moderate or strong impairment were included, although the prevalence was lower **(see appendix, figure S4)**. The self-reported fatigue as symptom cluster with its grades of interference with daily life correlated well with the standardised FAS questionnaire scores **(see appendix, table S1)**.

**Figure 2.**
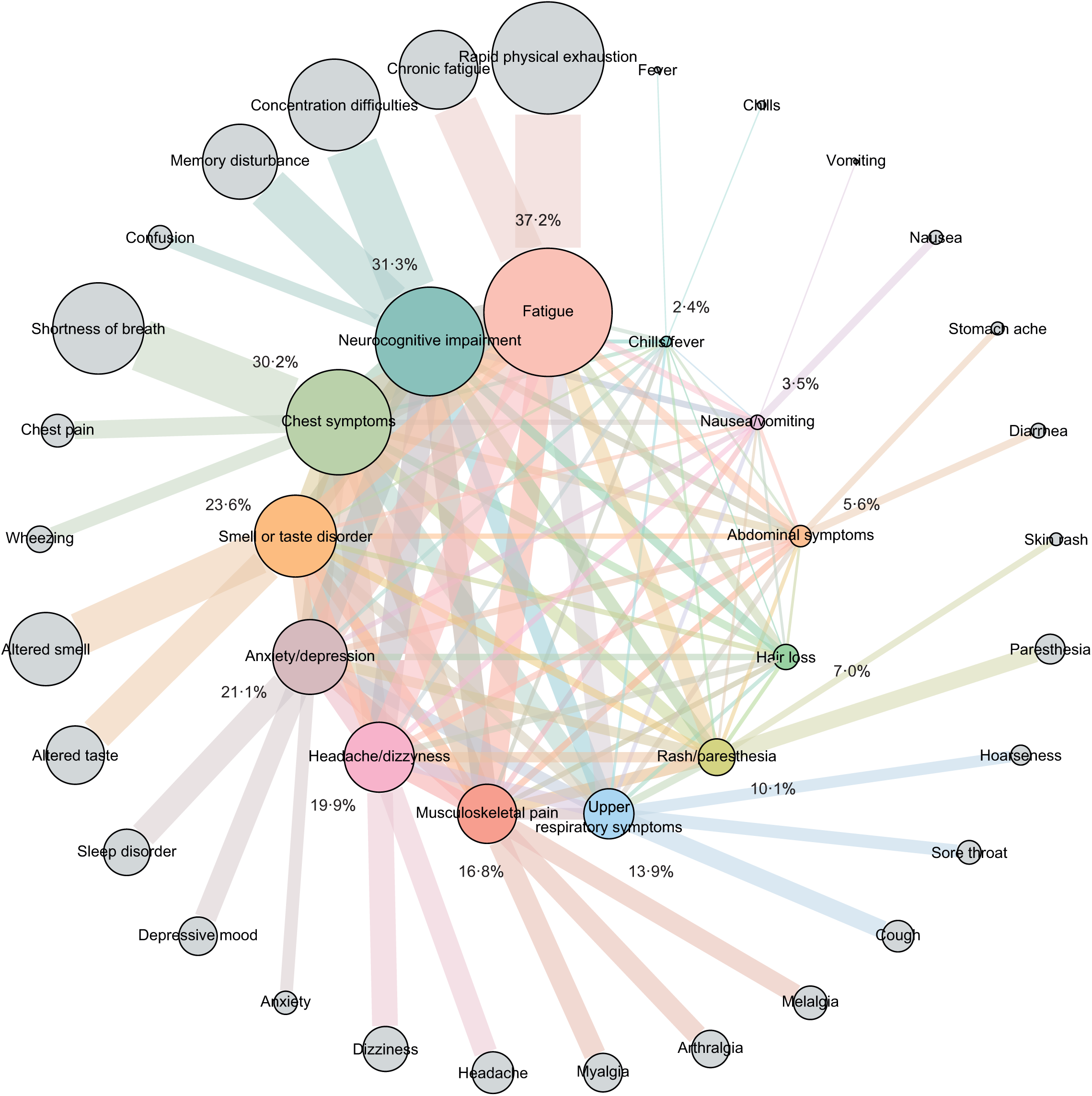
Co-occurrence network of symptom clusters 6-12 months after acute infection (with prevalence in %) along with contributing individual symptoms (outer circle). Only symptoms not present before the acute SARS-CoV-2 infection were considered.

We also looked at co-occurrence patterns between clusters. Interestingly, we found that smell or taste disorder was the cluster with the weakest co-occurrence with any other symptom cluster **(see appendix, figure S5)**, while fatigue as the most prevalent symptom cluster frequently co-occurred with neurocognitive impairment and chest symptoms.

### Associations of sociodemographic and other variables with symptom clusters

We explored determinants for the 13 symptom clusters **(see appendix, table S2)**. The mutually adjusted models included demographic and lifestyle variables, the severity of acute infection, time since infection and pre-existing comorbidities. Importantly, time since acute infection showed no association with symptom clusters (except for a weak association with an altered sense of smell/taste). The strongest consistent association was observed for initial out- or in-patient care versus no medical care during acute infection (as a proxy for severity of the initial infection), in particular for rash/paresthesia, chills/fever, and hair loss. The second strongest consistent determinant was female sex. Most of these associations became stronger when the analysis was restricted to symptom clusters with a grade of impairment moderate to strong **(see appendix, table S3)**.

BMI and smoking (particularly current smoker status) also appeared to be risk factors for several symptom clusters. Increasing age was a risk factor for fatigue, neurocognitive impairment and musculoskeletal pain (among others). Musculoskeletal and mental pre-existing disorders were associated with occurrence of reporting any symptom and with many different symptom clusters, while the associations of other pre-existing conditions with any or specific symptom clusters were variable and often weak.

### Impaired recovery of general health and working capacity

We next examined the association between symptom clusters and general health and working capacity (percentage recovered compared to prior acute infection). The self-reported mean health recovery was 89·5% (corresponding to an overall loss of 11·5%, 95% CI 11·2-11·7), and the overall loss of working capacity was 10·7% (95% CI 10·4-11·0), respectively. The various symptom clusters differed with regard to the associated loss of health and working capacity (**figure 3**). In terms of population-attributed loss, the fatigue cluster with the highest prevalence contributed most, with a 2·27% loss of general health (95% CI 2·07%-2·47%) and a loss of 2·32 % of working capacity (95% CI 2·09-2·56) – population attributable loss estimates for all other clusters were below 2%. Neurocognitive impairment had a significantly stronger effect on loss of working capacity than on loss of health. The opposite was found for chest symptoms and distorted sense of smell or taste, which both primarily affected general health recovery rather than working capacity **(figure 3)**. Again, there were notable differences according to age and sex **(see appendix, figure S6)**.

**Figure 3.**
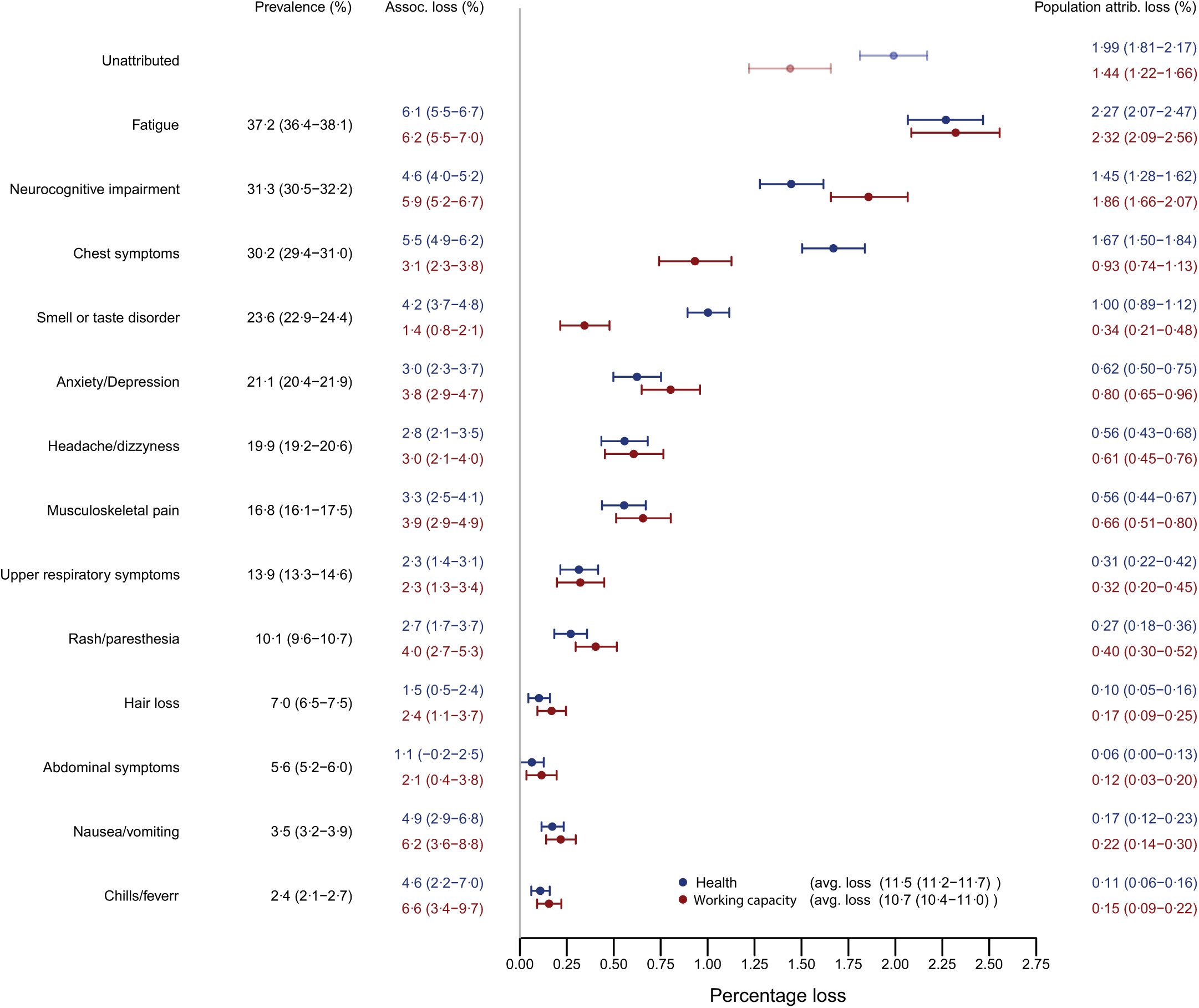
Prevalence of symptom clusters 6-12 months after acute infection (only symptoms not present before the acute SARS-CoV-2 infection) and associated loss (in %) and population attributable loss (in %) of general health and working capacity with 95% CI in brackets.

We finally examined how HRQoL correlated with health recovery and working capacity. We found a good correlation between the SF-12 physical subscore (but less so between the SF-12 mental health subscore) and both, health recovery (r=0·68) and working capacity (r=0·69) **(see appendix, figure S7)**.

Since functional consequences such as impaired health recovery or reduced working capacity might become key in estimating and discussing prevalence and burden of PCS among adults, we explored several scenarios for possible alternative case definitions. A shown in **figure 4**, almost one-third of the respondents (30·4%) reported their health recovery to be ≤80%, and 26·6% of the respondents reported ≤80% working capacity recovered in comparison to the situation before acute infection. If such reduced health or working capacity was combined with reporting (any) new symptom of moderate or strong impairment of daily life, we estimated a prevalence of 28·5% (corresponding to an age and sex-standardised prevalence of 26·5%).

**Figure 4.**
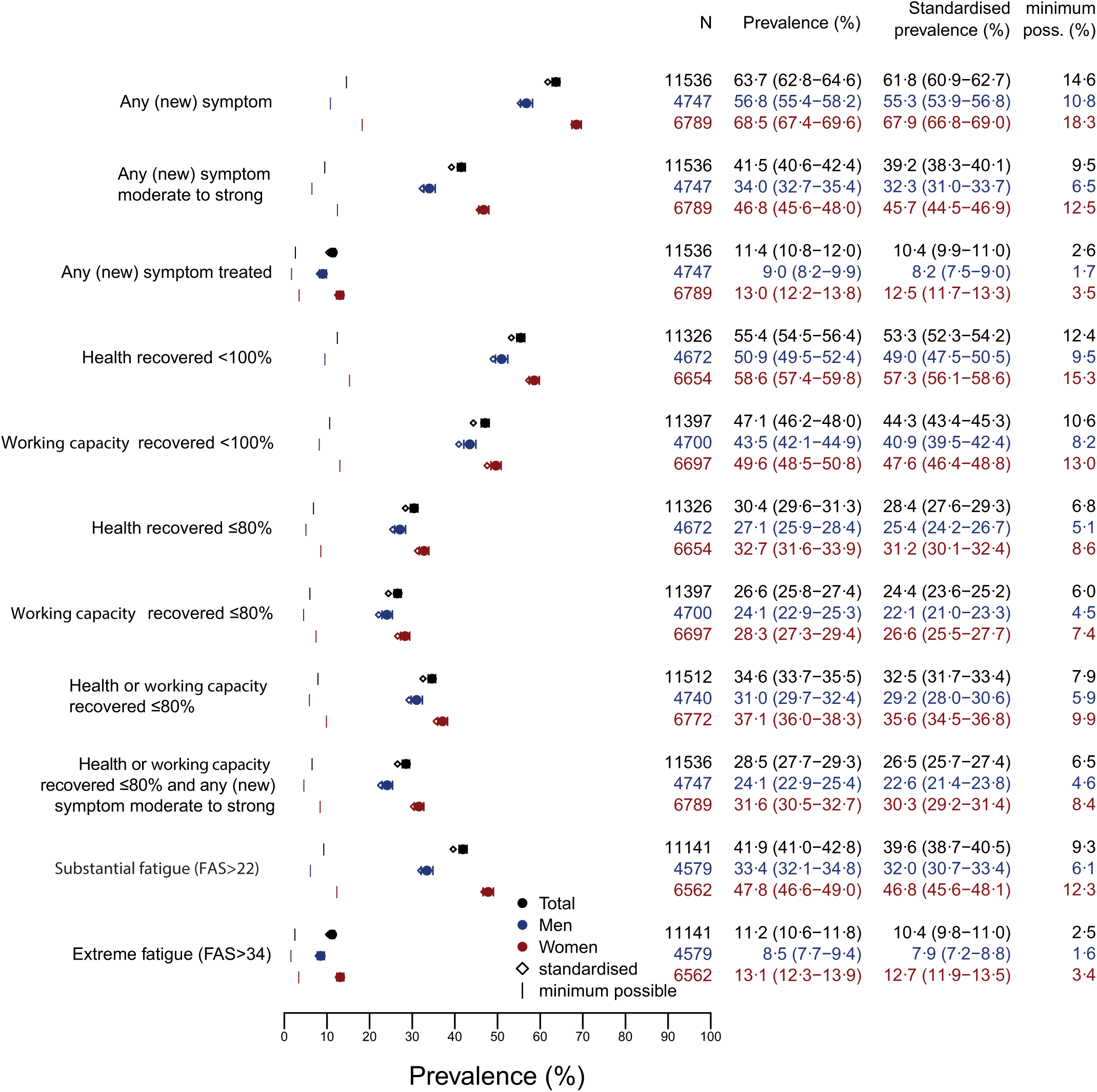
Prevalence (in %) of PCS according to different criteria for possible case definitions based on self-reported (new) symptoms, FAS (Fatigue Assessment score), recovered general health and working capacity.

## Discussion

This large population-based study found a considerable burden of symptoms with possible sequelae six to 12 months after SARS-CoV-2 infection affecting both general health and working capacity. Although a variety of long-lasting complaints was reported, few symptoms and symptom clusters drove this burden, and fatigue, neurocognitive impairment, and chest symptoms (e.g. shortness of breath) appeared to be the key health problems. The prevalence of post-acute symptoms remained substantial when considering only symptoms that had not been present before the acute infection or when considering only symptom clusters with moderate or strong degree of impairment, the latter resulting in estimated prevalence differences of 23% for fatigue, 15% for self-reported neurocognitive impairment, and 14% for chest symptoms, respectively, or a prevalence of 41·5% for any at least moderate symptom. A novel and important finding was that specific symptom clusters differed regarding their impact on health recovery and working capacity. Fatigue as well as neurocognitive impairment as the most prevalent health problems in this study also appeared to be most relevant for both impaired health recovery and reduced working capacity. Although chest symptoms were also associated with substantial loss of working capacity, their association with general health recovery remained comparably weak. A second important finding already observed by others was that most symptoms and symptom clusters were more frequent among women than among men and among individuals with more severe acute infection. Prevalence rates, however, appeared substantial among both men and women who had a mild course of acute SARS-CoV-2 infection, and PCS considerably affected also younger subjects. Unlike other studies, we limited our investigation to adults not older than 65 years, and this may explain why age was not a major determinant of symptom prevalence, but we show that age likely becomes relevant when regarding the functional consequences of the post-acute symptoms.

The prevalence of post-acute symptoms varies widely across studies.^6-9^ The available epidemiological study results have been challenging and difficult to interpret given the variety and heterogeneity of methodologies used, including the differences in selection of patient populations, availability of comparison groups, different follow-up periods, and inconsistent terms used to describe symptoms and adverse health conditions. Fatigue, respiratory and neurological sequelae, however, were the most frequent concerns in most previous work^13^, and the present study strongly suggests that this symptom complex affects even subjects with mild COVID-19 and younger populations more than half a year after acute infection.

The relevance in particular of fatigue and neurocognitive impairment is noteworthy for three reasons. First, fatigue or tiredness and exercise intolerance and similar problems are definitely more frequent in COVID-19 survivors than in control populations^14-20^ and have been the main complaints in many long covid studies, but few of them (12 of 43 evaluable studies in a recent review) used standardised instruments to quantify or validate self-reported symptoms of fatigue.^9^ The FAS instrument used by us and in a population-based Swiss study^15^ assesses fatigue largely distinct from depressive symptoms, anxiety, and neuroticism, and seemed to support the validity of self-reported symptoms of fatigue with different grades of impairment in our study. Whether alternative fatigue assessment instruments provide better sensitivity and specificity in the current pandemic setting is unknown. Second, fatigue was frequently accompanied by other prevalent symptom clusters such as chest pain and neurocognitive impairment, but also co-occurred with anxiety/depression as a symptom cluster including sleep disorders, and with many other complaints such as pain syndromes – similar to observations elsewhere.^14,21-24^ This may indicate some overlap of PCS with myalgic encephalomyelitis/chronic fatigue syndrome (ME/CFS) which may include similar sometimes relapsing symptoms and usually persists for years rather than for months. Further investigations are needed to address such a possible overlap.^25,26^ A third aspect is that neurocognitive impairment has not only frequently been self-reported after acute SARS-CoV-2 infection as in this study, but has already been validated in several studies as measurable deficiencies in reasoning, problem solving, spatial planning, target detection and diverse memory functions.^27-32^ At least some of the studies did not suggest improvement of cognitive performance measures after COVID-19 over time,^23,28^ and we also had no evidence of decreasing neurocognitive symptom prevalence within our observation period six to 12 months after acute infection. This may indicate that, similar to fatigue, this disorder might develop into a chronic health problem in an unknown proportion of patients.

The prevalence of post-acute symptoms has varied widely across studies also because often any symptom irrespective of whether already existing before COVID-19 or whether considered severe and functionally relevant has been included in interviews and questionnaires. In a large survey from the UK (with 76 155 subjects after confirmed acute SARS-CoV-2 infection), for example, self-reported tiredness and fatigue were quite frequent among the 37·7% subjects having (any) persistent symptoms 12 weeks or more after acute infection.^24^ However, only one third of the respondents considered their symptoms as being “severe”, the questionnaire did not include cognitive impairment items, and the number of respondents reporting one versus more than one symptom differed greatly, making a valid overall prevalence estimate of PCS difficult.

Estimating prevalence rates in our cohort based on different working definitions for PCS yielded a range between 64% (any post-acute symptom not present before acute infection included) and 11% (counting only extreme fatigue based on a correspondingly high FAS score) **(see figure 4)**. Others described a range of prevalences (at 12 weeks after acute infection) between 38% and 15% if (any) one or at least three symptoms were counted.^24^ Menges et al. in their population-based study found that roughly 25% of respondents had not fully recovered within six to eight months after SARS-CoV-2 infection – similar to the rate of roughly 30% found in the present study – but more than half of the survey participants reported symptoms of fatigue^15^ (when assessed by FAS) – slightly more than the 42% in the present study when using the same instrument. We believe that a PCS definition based on (any) reported symptom might easily overestimate PCS and become too non-specific if no functional or health-related quality of life or working capacity measure is taken into account. A proposal for an advanced working definition of PCS in our view might include the combination of symptoms with moderate or strong degree of impairment of daily life plus reduced (by at least 20%) health recovery or working capacity. Furthermore, clinical investigations using valid case definitions will be of great importance if mechanisms and risk factors of PCS including biomarkers are to be analysed in case-control studies.

Strengths of the present work are the large number of subjects, the defined period between six and 12 months after PCR-confirmed SARS-CoV-2 infection, and the population-based approach with inclusion of all infected subjects who were subject to the statutory reporting requirement within defined geographic regions. Furthermore, we used a within-subject comparison considering the symptom frequency before acute infection. Besides general health and working capacity, we included other measures assessing symptom severity and their individual as well as potential societal consequences such as work ability.

Limitations include the self-reported nature of symptoms and sequelae without medical validation. Also, reporting bias has to be considered when reporting symptoms from the past, especially in subjects with neurocognitive sequelae. Furthermore, we had a limited response with some overrepresentation of older persons and female sex **(appendix, table S4)**. Our study regions were located around medium sized university cities, with respondents having higher education than the general population, which may limit generalizability. As we only have a before-after comparison within infected subjects, we cannot differentiate between the impact of the pandemic itself and its consequences such as non-pharmaceutical and public health interventions on symptoms and symptom reporting from direct consequences of the virus infection. Finally, we used only one specific method for symptom clustering and cannot exclude that other methods would define different and presumably larger clusters.

## Conclusions

As one of the largest population-based studies with a follow-up of six to 12 months after acute SARS-CoV-2 infection, we show a considerable burden of symptoms with possible individual and societal relevant sequelae affecting also younger adults with a history of mild acute infection. Concentration difficulties or memory problems, shortness of breath, chronic fatigue, and rapid physical exhaustion showed an excess prevalence of more than 20% in the post-acute phase and considerably impaired general health and working capacity. Roughly one out of four patients had new symptoms that at least moderately impaired daily life and activities and were associated with reduced health recovery or working capacity. Given the individual and societal burden of post-COVID sequelae, the underlying biologic abnormalities and causes need urgent clarification to define adequate treatment options and develop effective rehabilitation measures.

## Data Availability

All data produced in the present study are available upon reasonable request to the authors.

## Acknowledgements

The authors thank all participants who took part in the survey. The authors acknowledge the participating local health authorities for their administrative and technical support. We thank key collaborators on this work: Nelli Edel, Bettina Deibert, Stefanie Döbele, Sabine Gerbersdorf, Katja Hirth, Achim Jerg, Moritz Munk, Sylvia Parthé, Stephan Rusch, Cynthia Stapornwongkul, Michaela Schmid, Patrick Roling, Jennifer Müller, Annika Noghero, and Hanna Tschischka.

