## Supplementary material for "Prevalence, determinants, and impact on general health and working capacity of post-acute sequelae of COVID-19 six to 12 months after infection: a population-based retrospective cohort study from southern Germany"

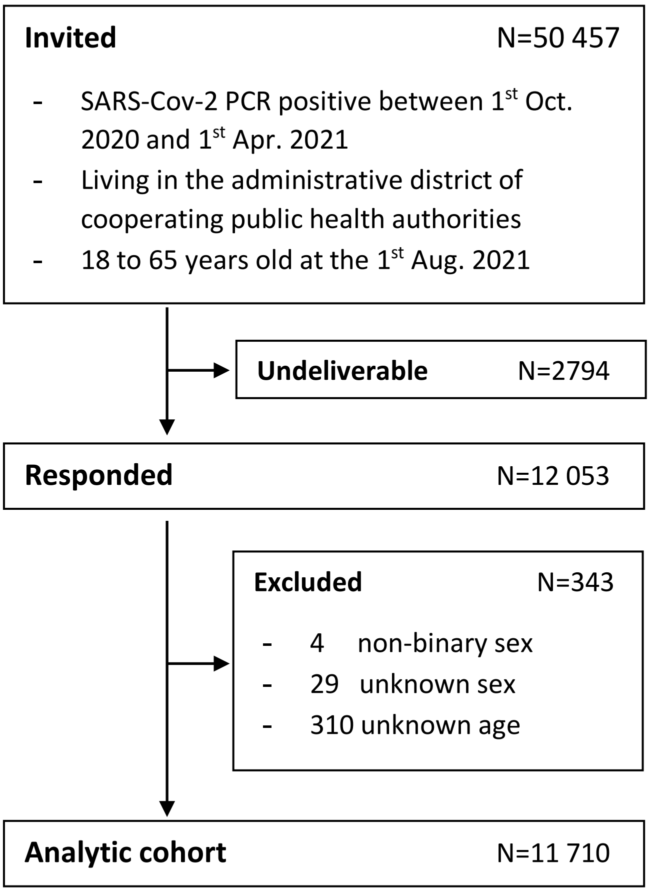

**Figure S1. Flow chart for the inclusion of SARS-CoV-2 PCR-positive subjects (tested between 1st October 2020 and 1st April 2021) aged 18-65 years from defined regions in southern Germany.**

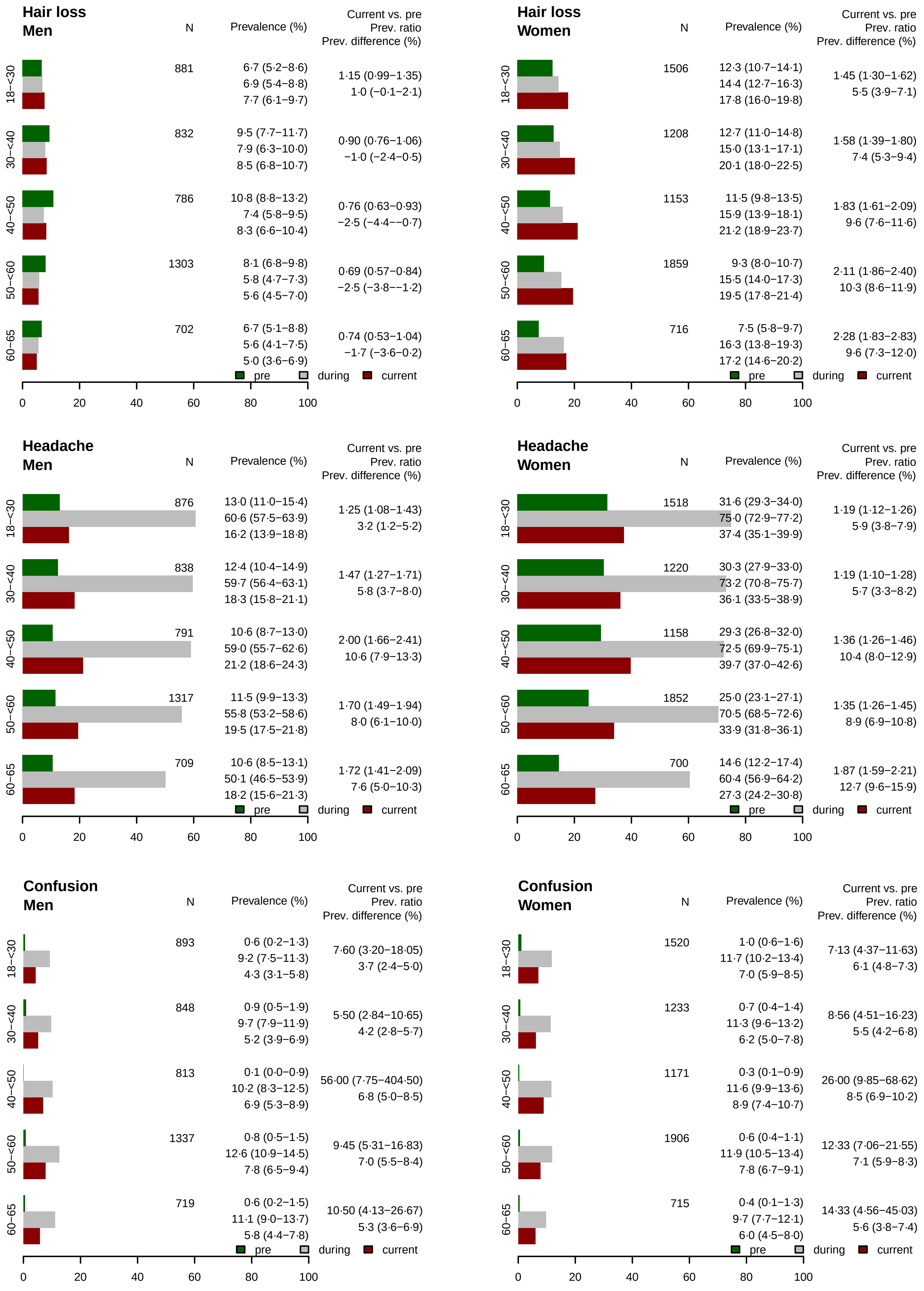

**Figure S2. Prevalence of symptoms before (pre), during, and 6-12 months after (current) after the SARS-CoV-2 infection, including prevalence ratio (current divided by pre) and prevalence difference (current minus pre) with 95 % confidence intervals according to age categories and sex. N represents the number of respondents in the respective age strata.**

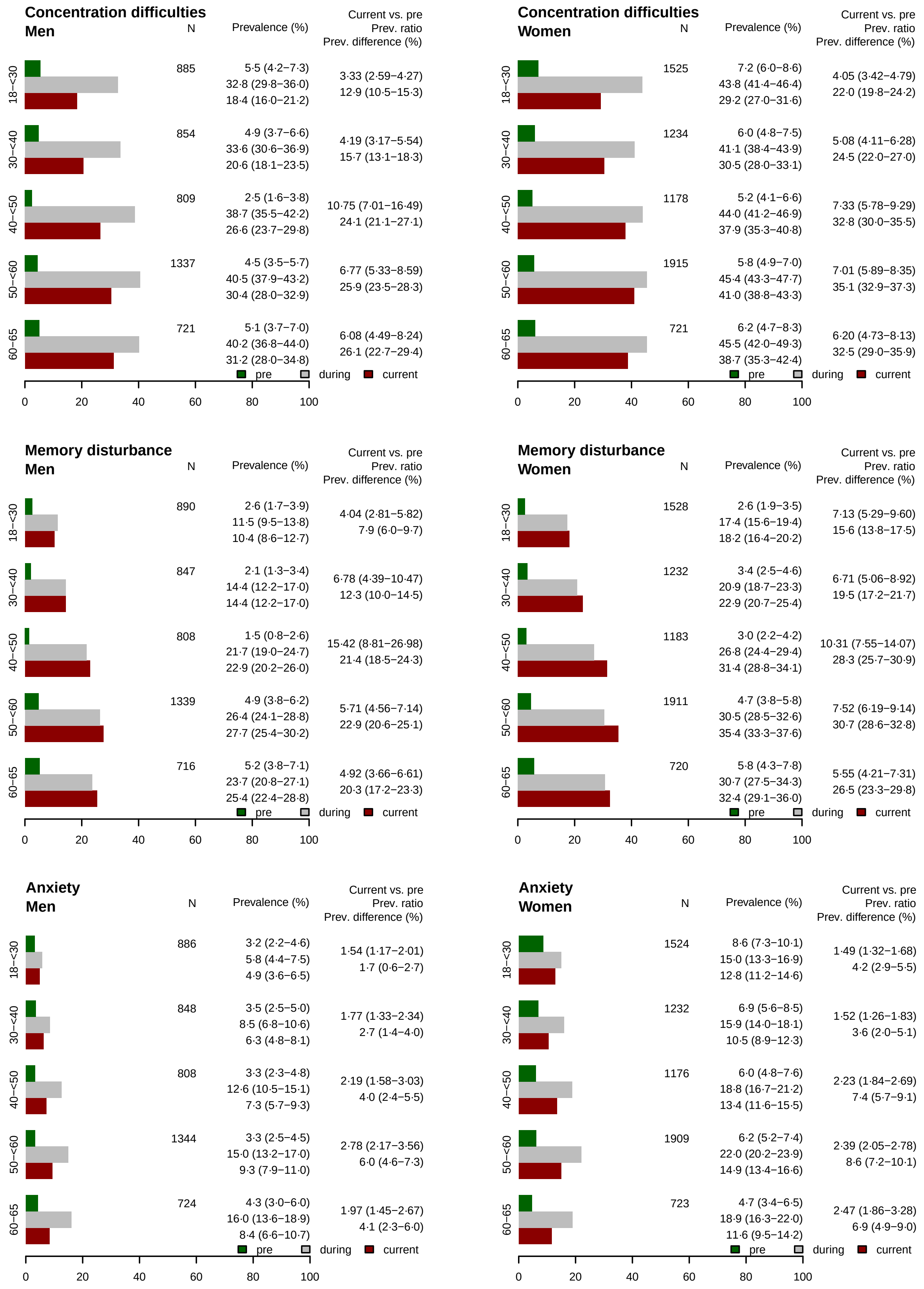

**Figure S2. Prevalence of symptoms before (pre), during, and 6-12 months after (current) after the SARS-CoV-2 infection, including prevalence ratio (current divided by pre) and prevalence difference (current minus pre) with 95 % confidence intervals according to age categories and sex. N represents the number of respondents in the respective age strata. (…continued)**

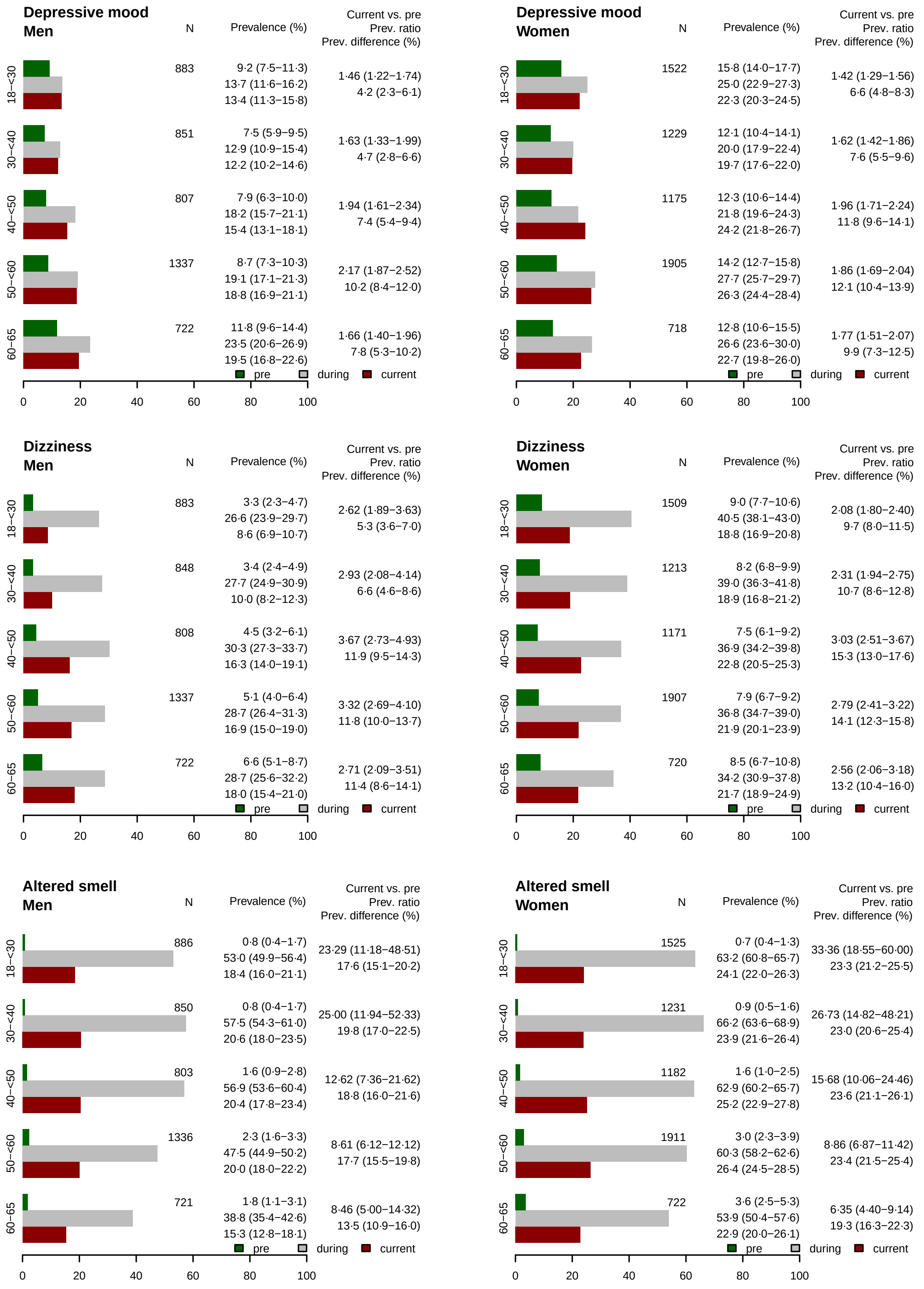

**Figure S2. Prevalence of symptoms before (pre), during, and 6-12 months after (current) after the SARS-CoV-2 infection, including prevalence ratio (current divided by pre) and prevalence difference (current minus pre) with 95 % confidence intervals according to age categories and sex. N represents the number of respondents in the respective age strata. (…continued)**

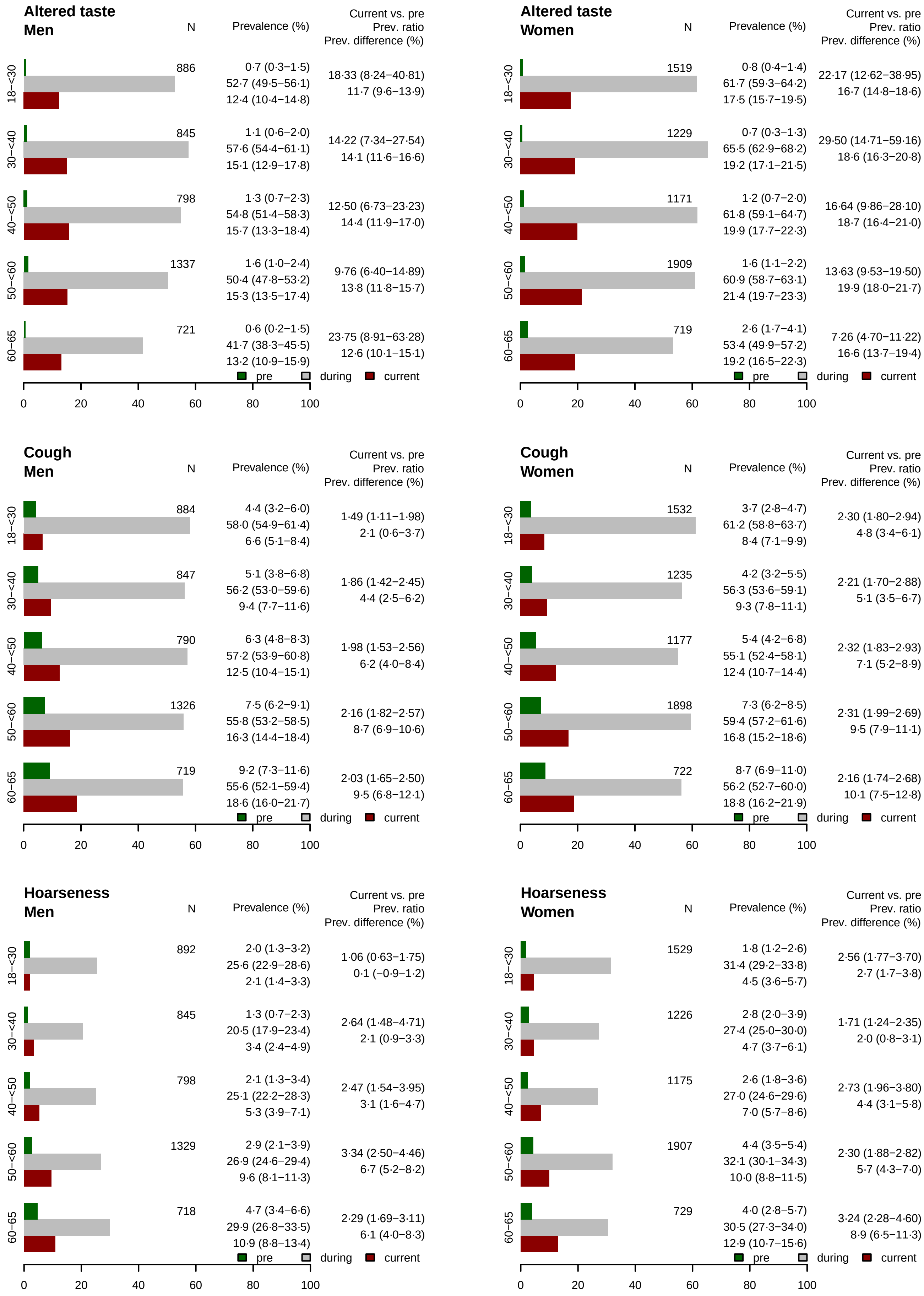

**Figure S2. Prevalence of symptoms before (pre), during, and 6-12 months after (current) after the SARS-CoV-2 infection, including prevalence ratio (current divided by pre) and prevalence difference (current minus pre) with 95 % confidence intervals according to age categories and sex. N represents the number of respondents in the respective age strata. (…continued)**

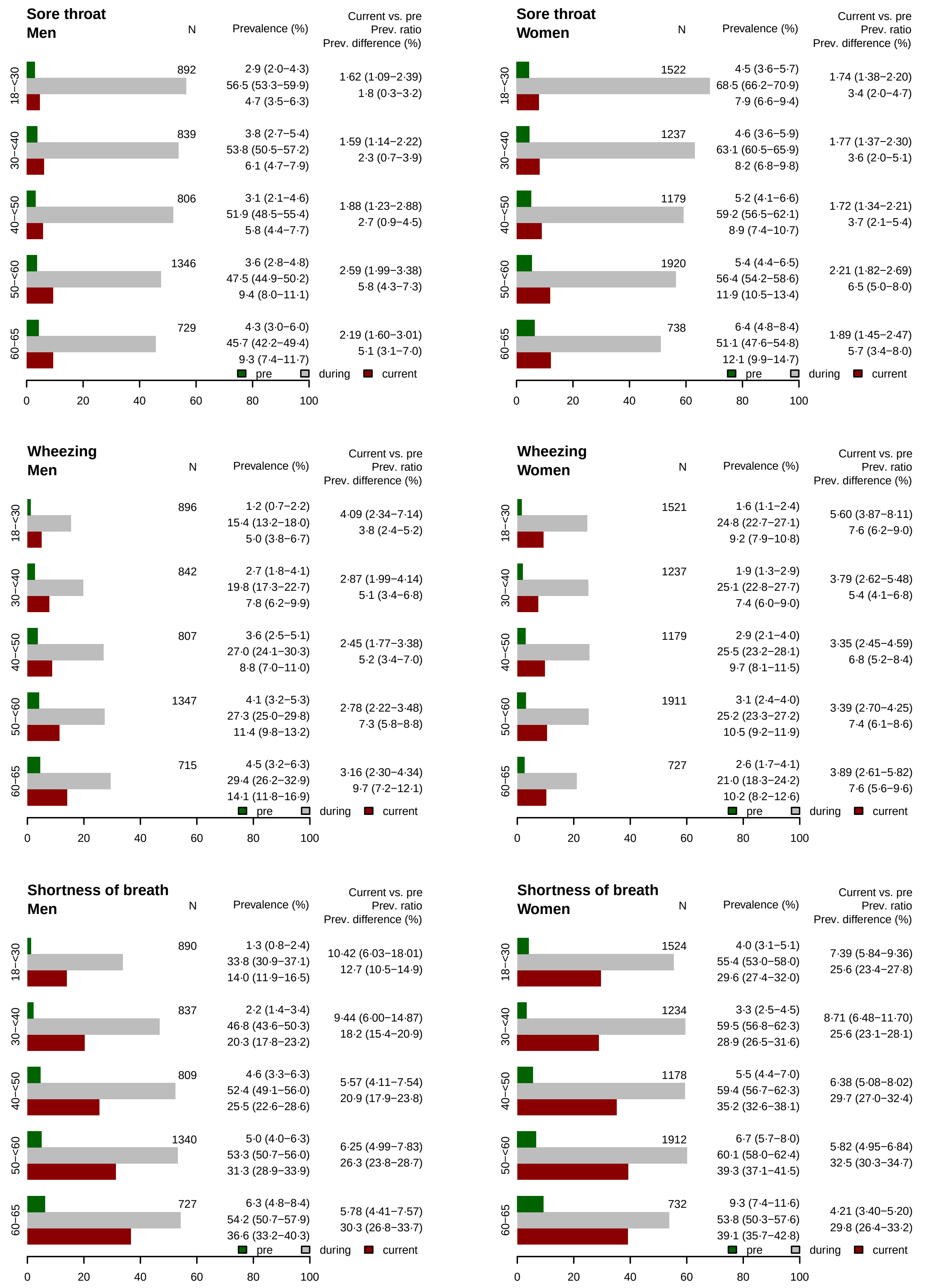

**Figure S2. Prevalence of symptoms before (pre), during, and 6-12 months after (current) after the SARS-CoV-2 infection, including prevalence ratio (current divided by pre) and prevalence difference (current minus pre) with 95 % confidence intervals according to age categories and sex. N represents the number of respondents in the respective age strata. (…continued)**

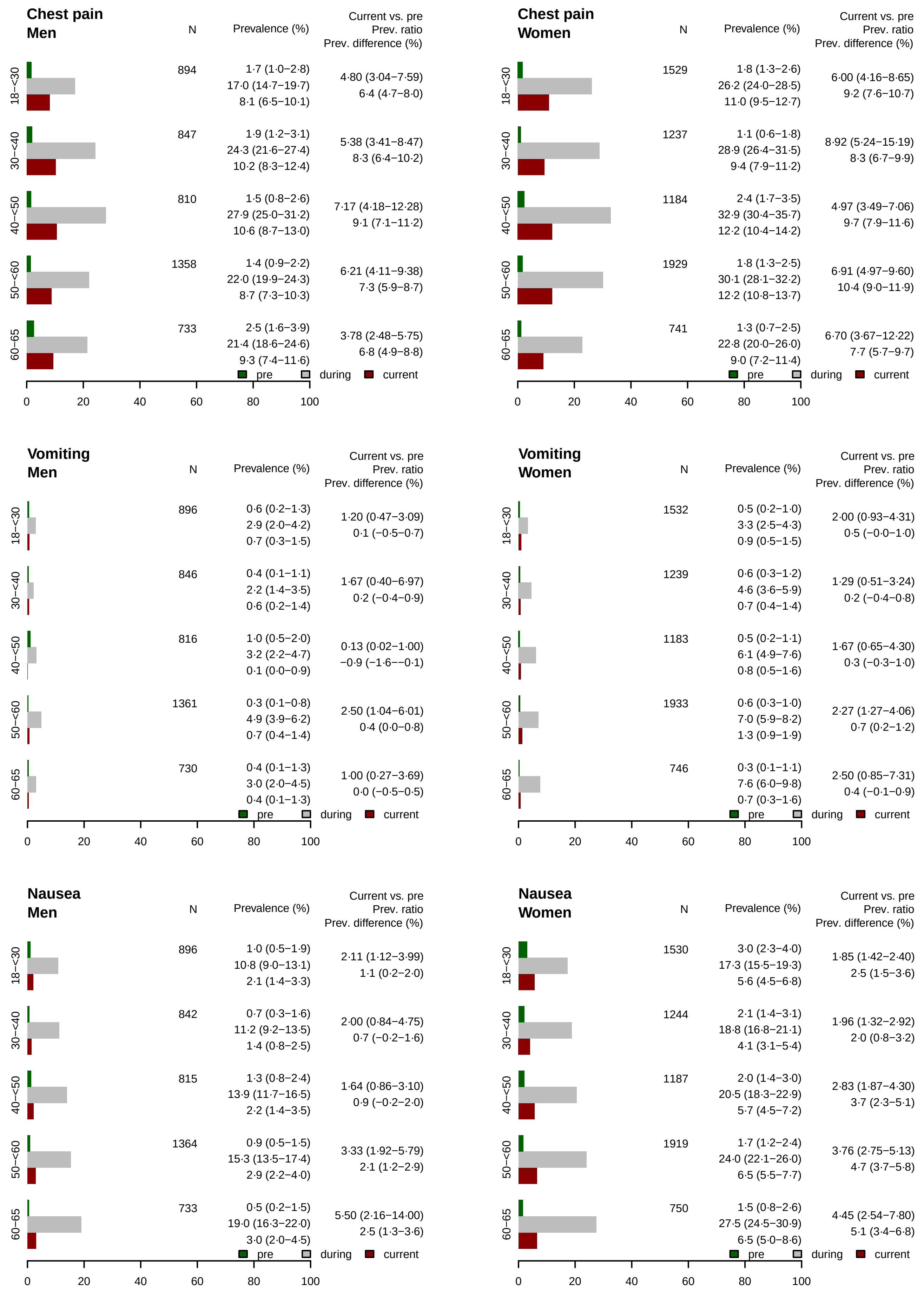

**Figure S2. Prevalence of symptoms before (pre), during, and 6-12 months after (current) after the SARS-CoV-2 infection, including prevalence ratio (current divided by pre) and prevalence difference (current minus pre) with 95 % confidence intervals according to age categories and sex. N represents the number of respondents in the respective age strata. (…continued)**

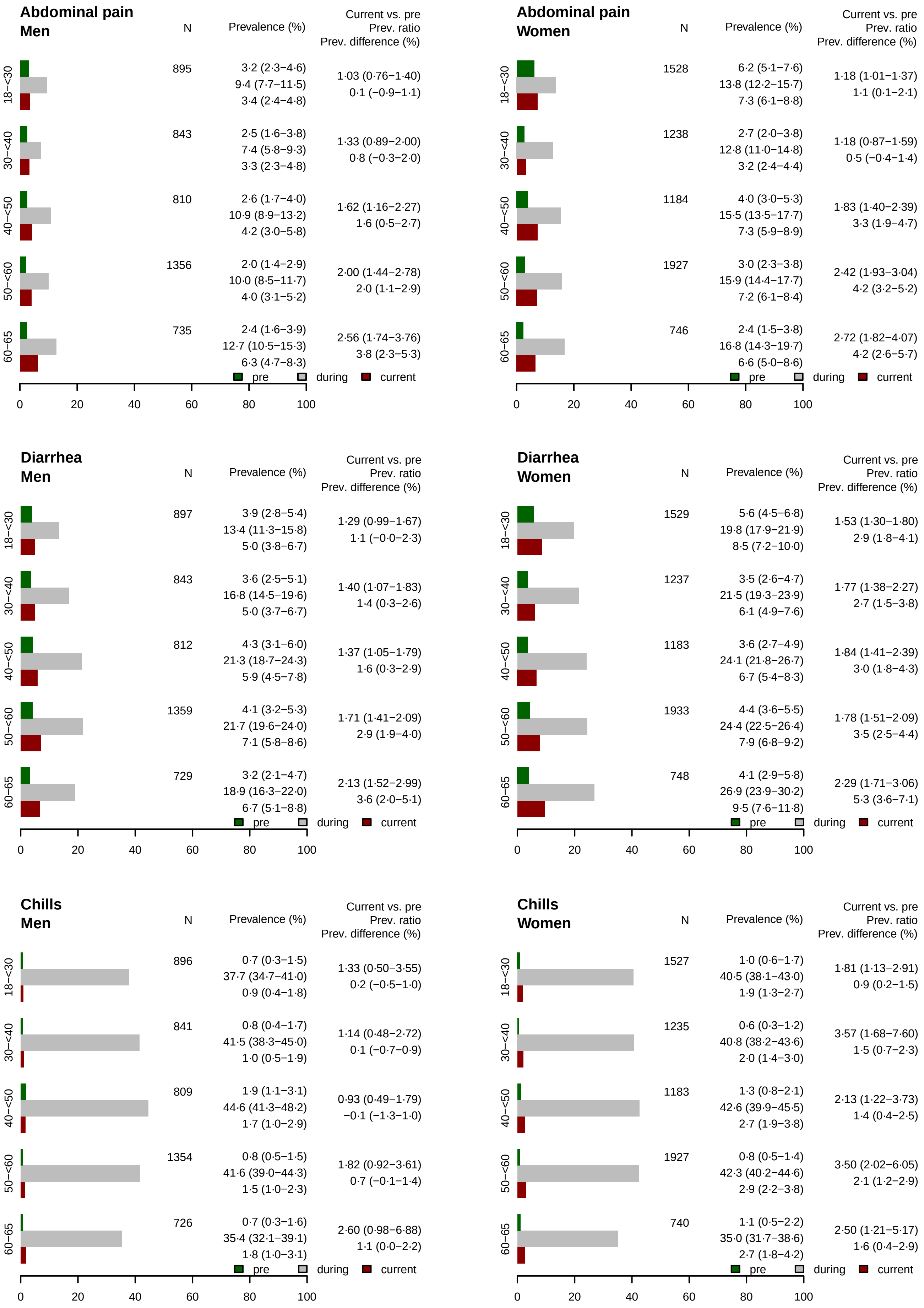

**Figure S2. Prevalence of symptoms before (pre), during, and 6-12 months after (current) after the SARS-CoV-2 infection, including prevalence ratio (current divided by pre) and prevalence difference (current minus pre) with 95 % confidence intervals according to age categories and sex. N represents the number of respondents in the respective age strata. (…continued)**

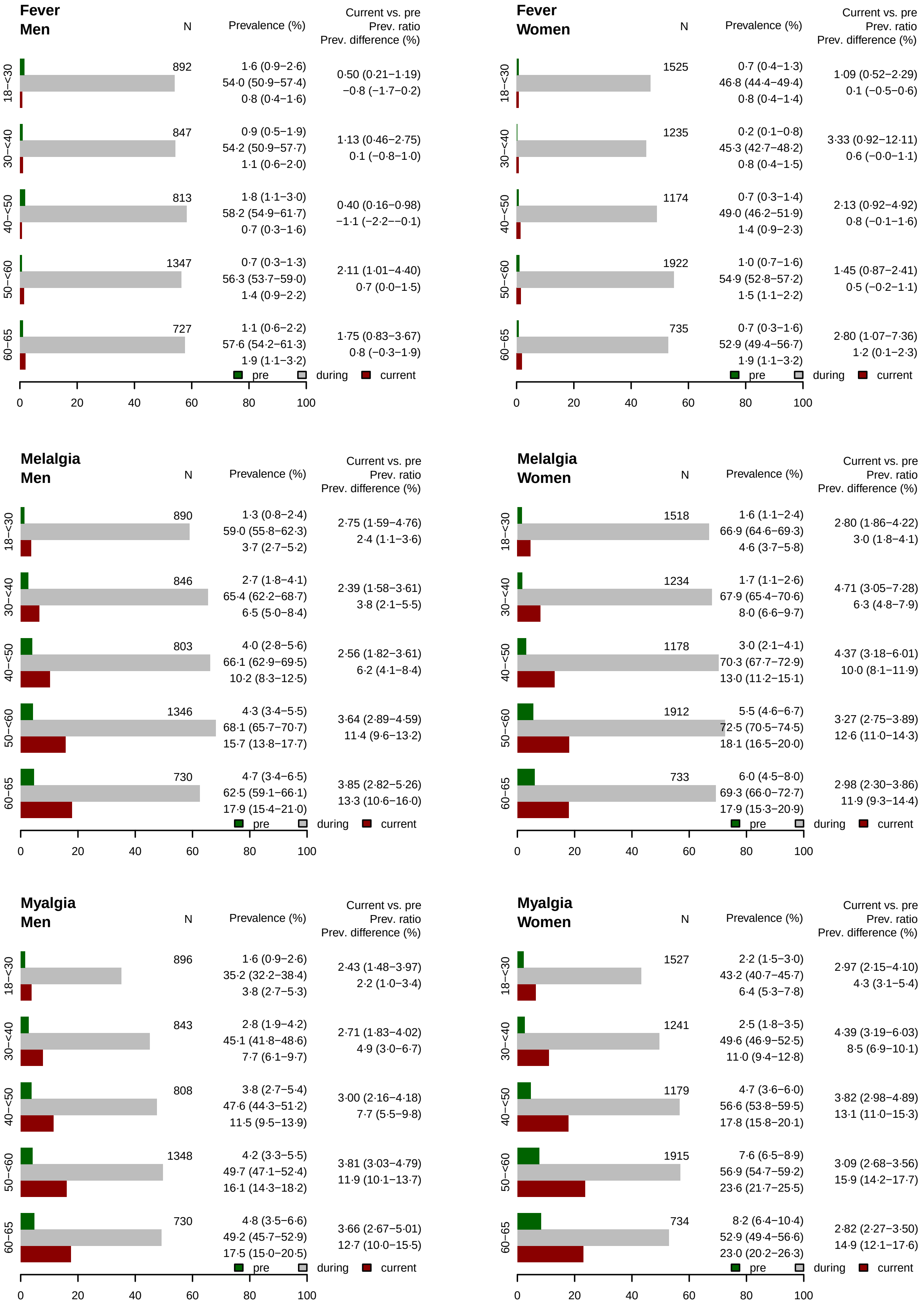

**Figure S2. Prevalence of symptoms before (pre), during, and 6-12 months after (current) after the SARS-CoV-2 infection, including prevalence ratio (current divided by pre) and prevalence difference (current minus pre) with 95 % confidence intervals according to age categories and sex. N represents the number of respondents in the respective age strata. (…continued)**

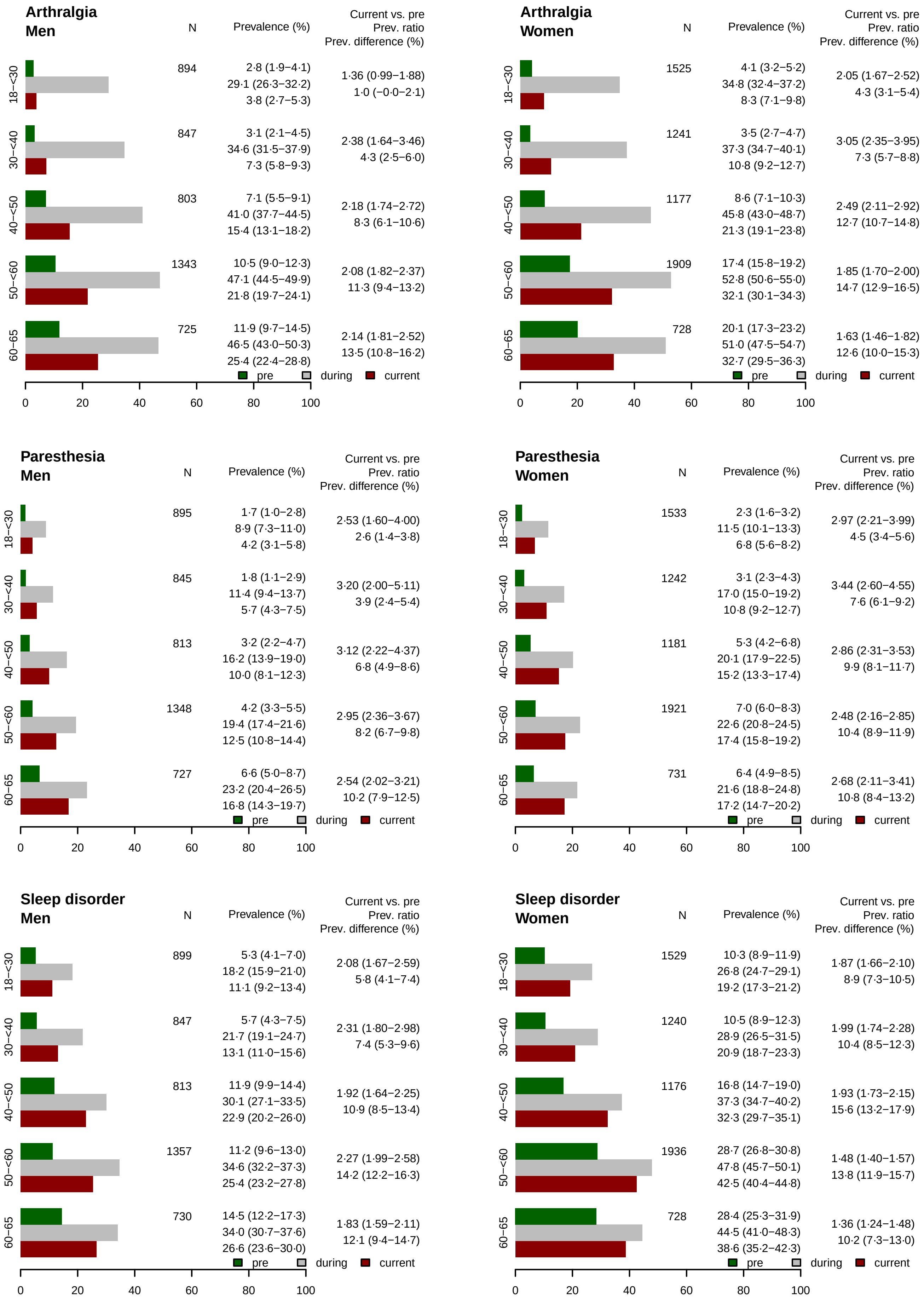

**Figure S2. Prevalence of symptoms before (pre), during, and 6-12 months after (current) after the SARS-CoV-2 infection, including prevalence ratio (current divided by pre) and prevalence difference (current minus pre) with 95 % confidence intervals according to age categories and sex. N represents the number of respondents in the respective age strata. (…continued)**

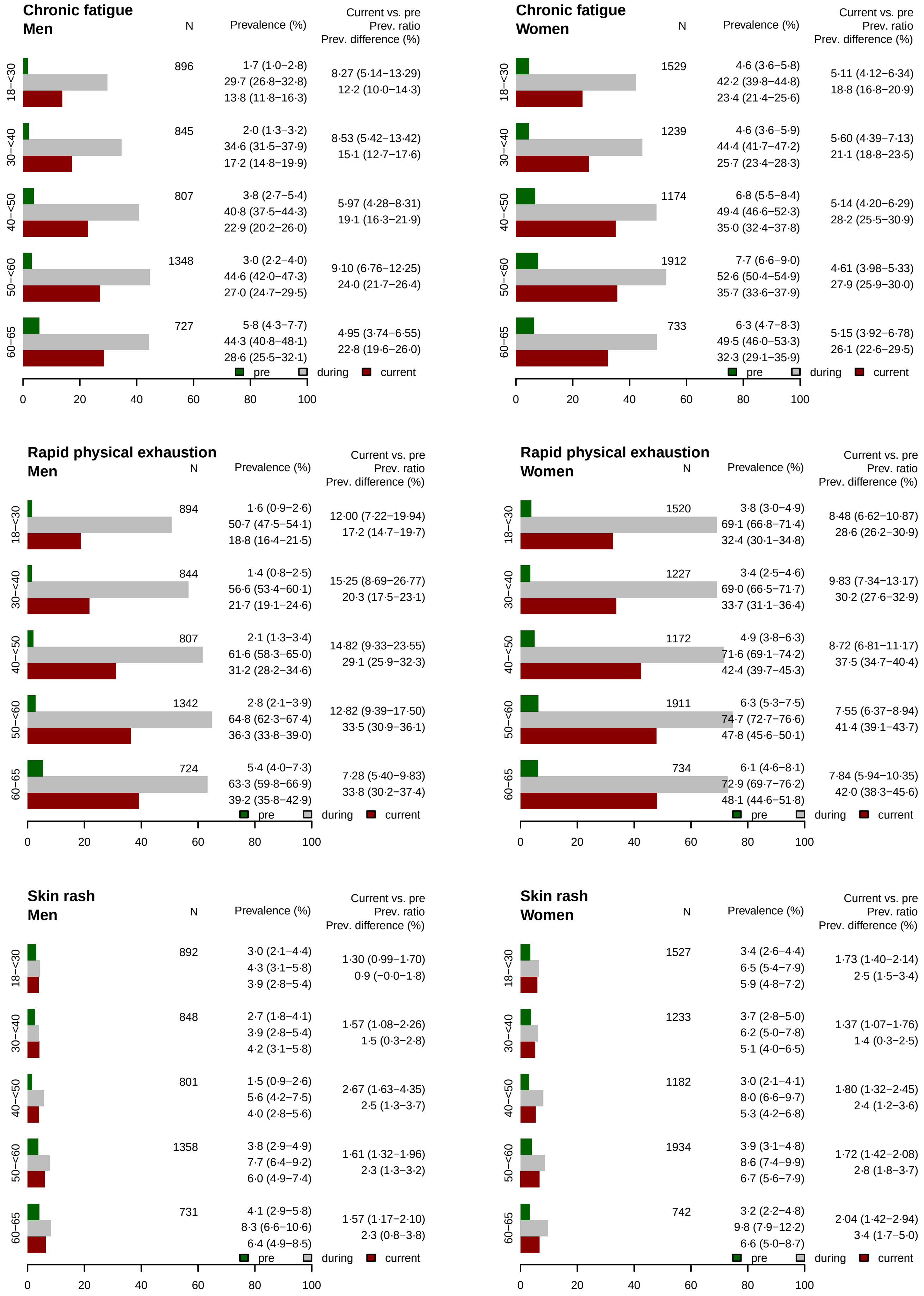

**Figure S2. Prevalence of symptoms before (pre), during, and 6-12 months after (current) after the SARS-CoV-2 infection, including prevalence ratio (current divided by pre) and prevalence difference (current minus pre) with 95 % confidence intervals according to age categories and sex. N represents the number of respondents in the respective age strata. (…continued)**

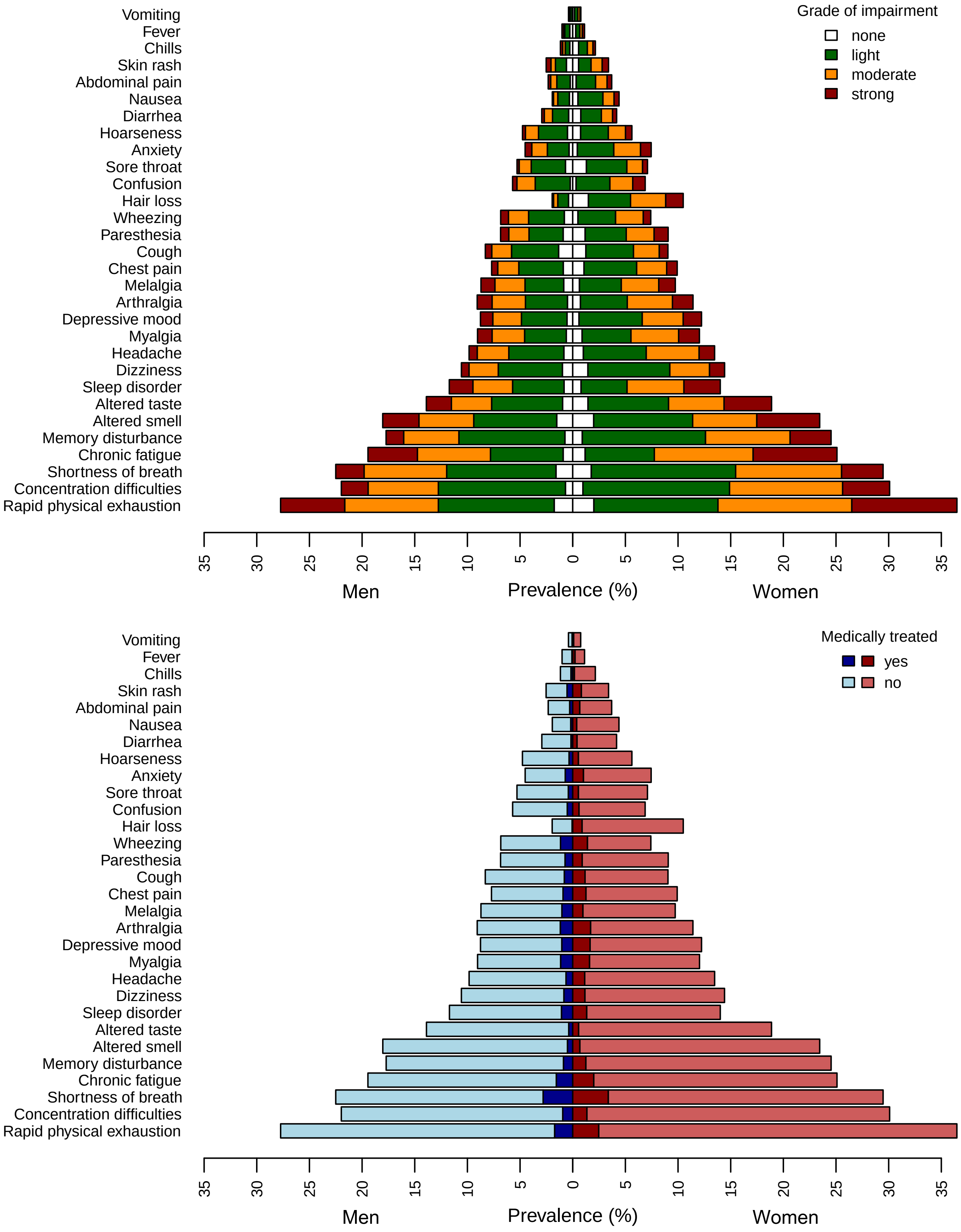

**Figure S3. Frequency of symptoms 6-12 months after acute infection, given the symptom was not present before the infection, stratified according to sex with grade of impairment (top panel) and medical treatment (bottom panel).**

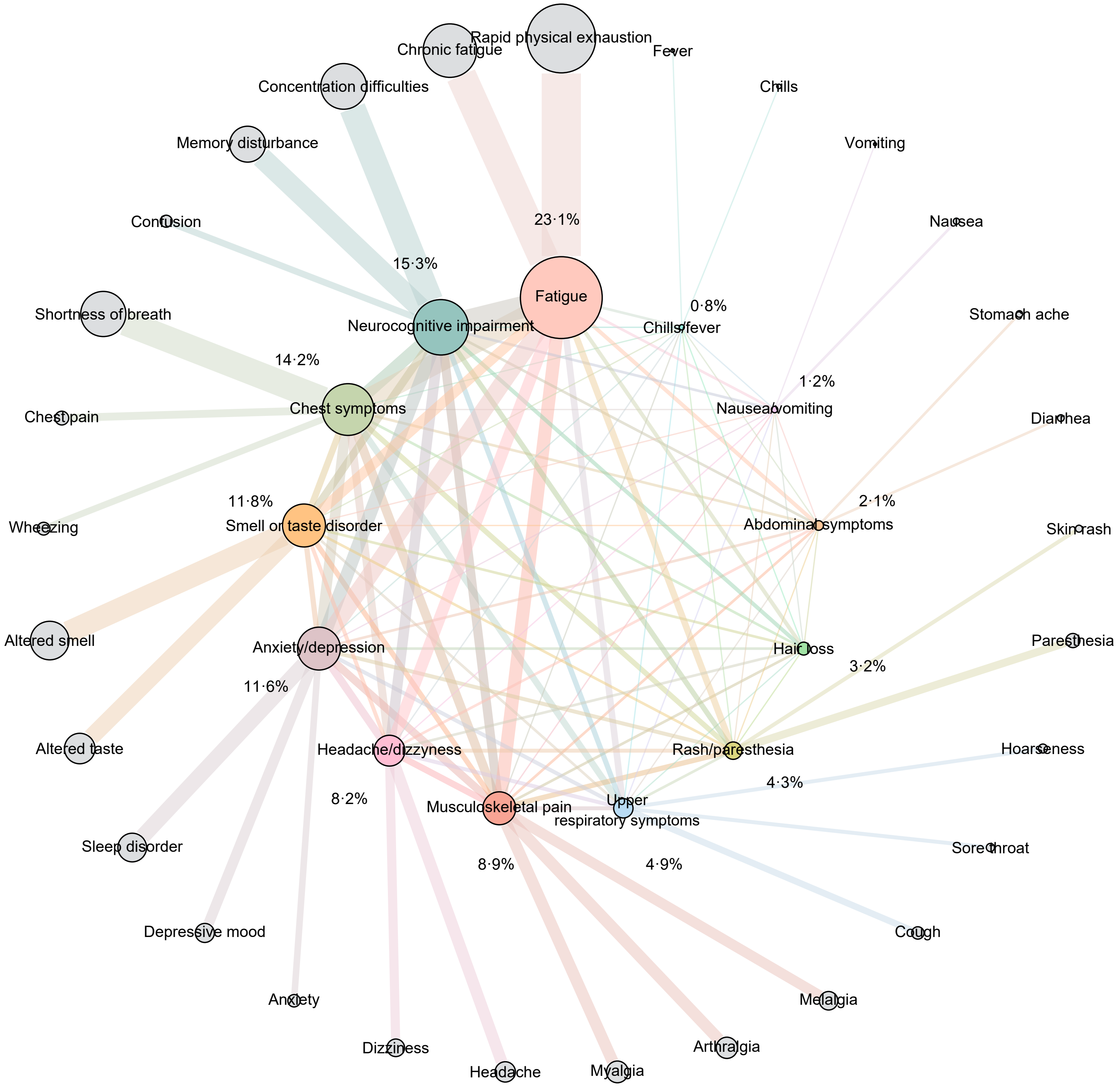

**Figure S4. Co-occurrence network of symptom clusters 6-12 months after SARS-CoV-2 infection (with prevalence in %), given the symptoms were not present before the infection, along with contributing individual symptoms of grade moderate to strong.**

**Table S1. Fatigue assessment scale (FAS) scores by new (not present before the infection) self-reported fatigue (as most prevalent symptom cluster)**

| Fatigue assessment scale (FAS) | All N=10 956 | No fatigue N=6879 | Fatigue with grade of impairment none/light N=1554 | moderate N=1443 | strong N=1080 |
|---|---|---|---|---|---|
| Total score, mean (sd) | 21·7 (8·9) | 17·7 (6.5) | 23·5 (6·4) | 28·7 (6·9) | 35·5 (7·3) |
| Physical sub-scale, mean (sd) | 11·9 (4·9) | 9·7 (3.7) | 13·0 (3·4) | 15·9 (3·5) | 19·3 (3·3) |
| Mental sub-scale, mean (sd) | 9·8 (4·5) | 8·0 (3.3) | 10·6 (3·6) | 12·7 (4·1) | 16·1 (4·7) |
| Proportion with FAS score >22, N (%) | 4570 (41·7) | 1409 (20·5) | 910 (58·6) | 1208 (83·7) | 1043 (96·6) |
| Proportion with FAS score >34, N (%) | 1228 (11·2) | 211 (3·1) | 95 (6·1) | 301 (20·9) | 621 (57·5) |

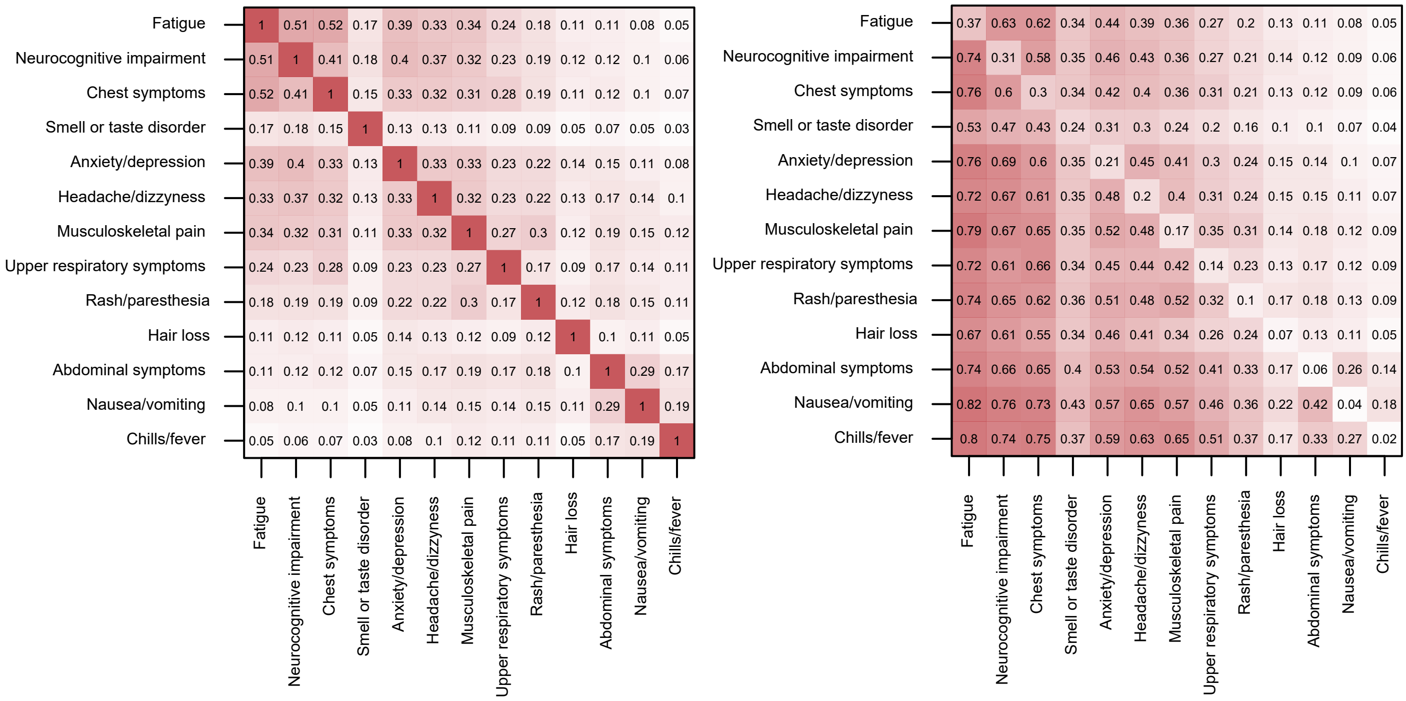

**Figure S5. Pairwise agreement (Cohen's kappa) between symptom clusters (left panel) and conditional probability (right panel) of experiencing symptoms of a specific cluster (columns) given another specific symptom cluster (rows). The overall prevalence of a specific symptom cluster is included in diagonal cells.**

**Table S2. Mutually adjusted prevalence ratios with 95% confidence intervals for new symptom clusters**

| | Any | Fatigue | Neurocognitive impairment | Chest symptoms | Smell or taste disorder | Anxiety/ depression | Headache/ dizziness |
|---|---|---|---|---|---|---|---|
| Age, per 10 years | 1·00 (0·99-1·01) | **1·04** (1·01-1·06) | **1·04** (1·01-1·06) | 0·98 (0·96-1·00) | **0·94** (0·92-0·97) | **1·05** (1·01-1·08) | 0·99 (0·96-1·02) |
| Sex | | | | | | | |
| Male (ref.) | | | | | | | |
| Female | **1·20** (1·16-1·24) | **1·27** (1·21-1·34) | **1·32** (1·24-1·40) | **1·27** (1·19-1·35) | **1·25** (1·16-1·35) | **1·32** (1·22-1·44) | **1·32** (1·21-1·44) |
| University entrance qualification | | | | | | | |
| Yes (ref.) | | | | | | | |
| No | 1·02 (0·99-1·06) | 1·05 (1·00-1·11) | 1·02 (0·96-1·08) | **1·08** (1·02-1·15) | 1·03 (0·96-1·11) | 1·03 (0·96-1·12) | **1·11** (1·03-1·21) |
| Smoking status, N (%) | | | | | | | |
| Never (ref.) | | | | | | | |
| Former | **1·07** (1·04-1·11) | **1·10** (1·04-1·17) | **1·18** (1·11-1·25) | **1·14** (1·07-1·22) | 1·05 (0·97-1·14) | **1·11** (1·02-1·21) | **1·14** (1·04-1·24) |
| Current | **1·06** (1·01-1·11) | **1·17** (1·08-1·26) | **1·11** (1·01-1·22) | **1·17** (1·06-1·28) | 1·03 (0·92-1·16) | **1·22** (1·08-1·38) | **1·24** (1·09-1·40) |
| BMI, per 5 kg/m² | **1·02** (1·01-1·03) | **1·05** (1·03-1·07) | **1·04** (1·02-1·07) | **1·10** (1·07-1·13) | 1·00 (0·96-1·03) | 1·02 (0·99-1·06) | **1·04** (1·01-1·08) |
| Time since positive PCR, per month | 1·00 (0·99-1·01) | 1·00 (0·98-1·01) | 1·01 (0·99-1·03) | 1·00 (0·98-1·02) | **1·04** (1·02-1·07) | 1·00 (0·98-1·03) | 0·99 (0·97-1·02) |
| Treatment of acute SARS-CoV-2 infection | | | | | | | |
| No medical care (ref.) | | | | | | | |
| Outpatient care | **1·36** (1·33-1·40) | **1·79** (1·70-1·88) | **1·87** (1·76-1·98) | **1·83** (1·72-1·95) | **1·33** (1·22-1·44) | **2·05** (1·89-2·22) | **1·94** (1·78-2·11) |
| Inpatient care | **1·41** (1·34-1·48) | **1·89** (1·73-2·07) | **1·93** (1·74-2·14) | **1·96** (1·75-2·18) | **1·23** (1·02-1·48) | **2·17** (1·87-2·52) | **2·10** (1·79-2·46) |
| Pre-existing conditions | | | | | | | |
| Musculoskeletal disorders | **1·12** (1·09-1·16) | **1·17** (1·11-1·23) | **1·20** (1·13-1·27) | **1·24** (1·16-1·32) | **1·22** (1·13-1·32) | **1·24** (1·14-1·34) | **1·29** (1·18-1·40) |
| Cardiovascular disorders | **1·07** (1·03-1·11) | **1·11** (1·04-1·18) | 1·04 (0·97-1·12) | **1·14** (1·06-1·23) | 1·04 (0·94-1·15) | 1·05 (0·95-1·16) | 1·02 (0·92-1·13) |
| Neurological or sensory disorders | **1·04** (1·01-1·08) | 1·05 (0·99-1·12) | **1·12** (1·05-1·20) | **1·13** (1·05-1·21) | 1·06 (0·97-1·16) | 1·05 (0·95-1·15) | 0·99 (0·89-1·09) |
| Metabolic disorders | **1·05** (1·02-1·09) | **1·12** (1·05-1·19) | **1·12** (1·05-1·20) | **1·09** (1·02-1·17) | 1·07 (0·98-1·18) | 1·06 (0·96-1·16) | **1·13** (1·03-1·25) |
| Mental disorders | **1·10** (1·06-1·14) | **1·16** (1·09-1·24) | **1·19** (1·11-1·28) | **1·11** (1·02-1·19) | 1·08 (0·98-1·19) | 0·91 (0·81-1·01) | **1·17** (1·05-1·30) |
| Respiratory diseases | **1·06** (1·02-1·10) | **1·15** (1·08-1·22) | **1·12** (1·04-1·20) | **1·09** (1·01-1·18) | 0·91 (0·82-1·01) | 1·04 (0·94-1·15) | 1·10 (0·99-1·22) |
| Dermatological diseases | 1·03 (0·99-1·08) | 1·01 (0·94-1·08) | 1·04 (0·96-1·13) | 1·03 (0·95-1·12) | 1·07 (0·96-1·19) | 1·10 (0·98-1·22) | 0·92 (0·82-1·04) |
| Cancer | 1·02 (0·95-1·09) | 0·97 (0·86-1·10) | 1·03 (0·91-1·18) | 0·99 (0·85-1·14) | 1·08 (0·91-1·30) | 0·87 (0·71-1·07) | 0·95 (0·77-1·17) |

**Table S2. Mutually adjusted prevalence ratios with 95% confidence intervals for new symptom clusters (…continued)**

| | Musculoskeletal pain | Upper respiratory symptoms | Rash/paresthesia | Hair loss | Abdominal symptoms | Nausea/vomiting | Chills/fever |
|---|---|---|---|---|---|---|---|
| Age, per 10 years | **1·20** (1·16-1·25) | **1·12** (1·07-1·16) | **1·10** (1·04-1·15) | 0·97 (0·92-1·03) | 1·02 (0·95-1·09) | 1·01 (0·92-1·10) | 1·07 (0·95-1·20) |
| Sex | | | | | | | |
| Male (ref.) | | | | | | | |
| Female | **1·36** (1·24-1·49) | **1·14** (1·03-1·26) | **1·36** (1·20-1·54) | **4·77** (3·86-5·90) | **1·35** (1·13-1·61) | **2·02** (1·57-2·60) | **1·51** (1·13-2·01) |
| University entrance qualification | | | | | | | |
| Yes | | | | | | | |
| No | **1·13** (1·03-1·24) | **1·17** (1·06-1·30) | **0·88** (0·78-0·99) | 1·12 (0·97-1·30) | 1·17 (0·99-1·39) | **1·27** (1·02-1·58) | 1·14 (0·86-1·51) |
| Smoking status, N (%) | | | | | | | |
| Never (ref.) | | | | | | | |
| Former | 1·07 (0·97-1·18) | 0·93 (0·83-1·04) | **1·20** (1·05-1·37) | 0·99 (0·84-1·17) | 1·03 (0·86-1·24) | **1·37** (1·09-1·73) | 1·25 (0·93-1·66) |
| Current | **1·37** (1·20-1·56) | **1·28** (1·11-1·49) | **1·41** (1·17-1·70) | 1·02 (0·81-1·30) | 1·08 (0·83-1·42) | **1·60** (1·18-2·17) | **1·72** (1·17-2·51) |
| BMI, per 5 kg/m² | **1·08** (1·04-1·12) | **1·09** (1·04-1·14) | **1·08** (1·02-1·14) | **1·07** (1·01-1·13) | 0·98 (0·90-1·07) | 0·94 (0·84-1·05) | 1·01 (0·89-1·14) |
| Time since positive PCR, per month | 0·99 (0·97-1·02) | 0·98 (0·95-1·01) | 1·01 (0·98-1·05) | **0·88** (0·84-0·92) | 0·98 (0·93-1·03) | 0·96 (0·90-1·02) | 0·96 (0·88-1·03) |
| Treatment of acute SARS-CoV-2 infection | | | | | | | |
| No medical care (ref.) | | | | | | | |
| Outpatient care | **2·16** (1·97-2·37) | **2·07** (1·86-2·29) | **2·08** (1·82-2·37) | **2·24** (1·93-2·61) | **2·41** (2·02-2·88) | **2·33** (1·85-2·92) | **3·08** (2·33-4·06) |
| Inpatient care | **2·17** (1·83-2·56) | **1·79** (1·46-2·20) | **3·03** (2·45-3·74) | **4·06** (3·22-5·11) | **2·74** (1·98-3·78) | **2·70** (1·76-4·14) | **3·42** (2·10-5·57) |
| Pre-existing conditions | | | | | | | |
| Musculoskeletal disorders | **1·21** (1·10-1·32) | **1·29** (1·16-1·43) | 1·13 (0·99-1·29) | 1·07 (0·91-1·25) | **1·22** (1·02-1·46) | 1·23 (0·98-1·55) | 1·07 (0·81-1·41) |
| Cardiovascular disorders | **1·13** (1·02-1·25) | **1·15** (1·02-1·29) | 1·02 (0·87-1·19) | 1·17 (0·97-1·41) | 1·22 (1·00-1·49) | 1·01 (0·77-1·33) | 1·08 (0·79-1·49) |
| Neurological or sensory disorders | 1·10 (0·99-1·22) | **1·14** (1·01-1·28) | **1·25** (1·08-1·44) | 1·02 (0·85-1·23) | 1·03 (0·84-1·27) | 1·15 (0·89-1·47) | 0·88 (0·63-1·23) |
| Metabolic disorders | 1·09 (0·98-1·21) | 1·07 (0·95-1·21) | **1·17** (1·01-1·35) | **1·19** (1·01-1·41) | 1·19 (0·97-1·45) | 1·27 (0·99-1·62) | 1·08 (0·79-1·47) |
| Mental disorders | **1·18** (1·06-1·32) | 1·11 (0·97-1·26) | **1·18** (1·01-1·38) | 1·09 (0·90-1·32) | **1·31** (1·06-1·62) | **1·70** (1·33-2·18) | 1·21 (0·84-1·72) |
| Respiratory diseases | 1·05 (0·94-1·18) | **1·17** (1·03-1·33) | 1·16 (0·99-1·36) | 0·94 (0·76-1·14) | 0·99 (0·79-1·24) | 1·07 (0·81-1·42) | 1·18 (0·83-1·66) |
| Dermatological diseases | 0·97 (0·85-1·11) | 1·06 (0·92-1·22) | 1·17 (0·99-1·37) | 1·10 (0·90-1·35) | 0·89 (0·69-1·15) | 0·95 (0·70-1·29) | 1·17 (0·81-1·68) |
| Cancer | 1·13 (0·94-1·37) | 1·08 (0·86-1·34) | 0·81 (0·59-1·12) | 0·88 (0·62-1·27) | 1·10 (0·75-1·61) | 1·05 (0·65-1·71) | 1·27 (0·71-2·25) |

**Table S3. Mutually adjusted prevalence ratios with 95% confidence intervals for new symptom clusters with grade of impairment moderate to strong**

| | Any | Fatigue | Neurocognitive impairment | Chest symptoms | Smell or taste disorder | Anxiety/depression | Headache/ dizziness |
|---|---|---|---|---|---|---|---|
| Age, per 10 years | 1·00 (0·98-1·02) | **1·06** (1·02-1·09) | **1·05** (1·01-1·09) | 0·99 (0·95-1·04) | 0·96 (0·92-1·00) | 1·03 (0·99-1·08) | 0·99 (0·94-1·05) |
| Sex | | | | | | | |
| Male (ref.) | | | | | | | |
| Female | **1·37** (1·30-1·44) | **1·46** (1·35-1·58) | **1·52** (1·37-1·68) | **1·33** (1·20-1·48) | **1·36** (1·21-1·53) | **1·47** (1·30-1·66) | **1·49** (1·29-1·72) |
| University entrance qualification | | | | | | | |
| Yes (ref.) | | | | | | | |
| No | **1·14** (1·09-1·20) | **1·19** (1·10-1·28) | **1·09** (0·99-1·20) | **1·25** (1·13-1·39) | **1·10** (0·99-1·23) | **1·15** (1·02-1·29) | **1·37** (1·19-1·57) |
| Smoking status, N (%) | | | | | | | |
| Never (ref.) | | | | | | | |
| Former | **1·12** (1·06-1·17) | **1·12** (1·03-1·21) | **1·25** (1·13-1·38) | **1·18** (1·06-1·31) | 1·07 (0·94-1·21) | **1·24** (1·09-1·40) | 1·13 (0·97-1·31 |
| Current | **1·22** (1·14-1·31) | **1·37** (1·24-1·53) | **1·26** (1·09-1·46) | **1·27** (1·09-1·48) | **1·30** (1·11-1·54) | **1·49** (1·26-1·75) | **1·34** (1·10-1·64) |
| BMI, per 5 kg/m² | **1·05** (1·03-1·07) | **1·09** (1·05-1·12) | **1·06** (1·02-1·11) | **1·16** (1·11-1·20) | **1·08** (1·02-1·13) | 1·04 (0·99-1·10) | 1·04 (0·97-1·10) |
| Time since positive PCR, per month | 1·00 (0·98-1·01) | 0·98 (0·96-1·01) | 1·01 (0·98-1·03) | 1·01 (0·98-1·04) | **1·03** (1·00-1·07) | 1·00 (0·96-1·03) | 0·97 (0·94-1·02) |
| Treatment of acute SARS-CoV-2 infection | | | | | | | |
| No medical care (ref.) | | | | | | | |
| Outpatient care | **1·69** (1·62-1·77) | **2·12** (1·97-2·28) | **2·27** (2·05-2·50) | **2·43** (2·19-2·69) | **1·48** (1·31-1·68) | **2·50** (2·23-2·80) | **2·79** (2·42-3·21) |
| Inpatient care | **1·82** (1·68-1·97) | **2·18** (1·91-2·50) | **2·64** (2·24-3·11) | **2·52** (2·10-3·02) | **1·29** (0·98-1·71) | **2·57** (2·06-3·20) | **3·16** (2·46-4·05) |
| Pre-existing conditions | | | | | | | |
| Musculoskeletal disorders | **1·19** (1·14-1·25) | **1·23** (1·14-1·33) | **1·30** (1·18-1·43) | **1·32** (1·19-1·46) | **1·24** (1·10-1·40) | **1·18** (1·04-1·33) | **1·34** (1·17-1·55) |
| Cardiovascular disorders | **1·11** (1·05-1·17) | **1·12** (1·03-1·22) | 0·99 (0·88-1·11) | **1·12** (1·00-1·27) | 0·97 (0·83-1·12) | 1·02 (0·88-1·18) | 0·92 (0·77-1·10) |
| Respiratory diseases | **1·11** (1·05-1·18) | **1·20** (1·10-1·31) | **1·16** (1·03-1·30) | **1·19** (1·05-1·35) | 0·93 (0·79-1·09) | 1·03 (0·89-1·20) | 1·01 (0·84-1·21) |
| Mental disorders | **1·20** (1·14-1·27) | **1·26** (1·16-1·37) | **1·46** (1·31-1·63) | **1·17** (1·03-1·32) | 1·07 (0·92-1·24) | 1·04 (0·89-1·21) | **1·38** (1·17-1·62) |
| Neurological or sensory disorders | **1·09** (1·03-1·15) | **1·15** (1·06-1·25) | 1·11 (0·99-1·23) | **1·25** (1·12-1·40) | 1·11 (0·97-1·28) | 1·05 (0·91-1·20) | 0·97 (0·82-1·15) |
| Dermatological diseases | 1·05 (0·99-1·12) | 1·04 (0·94-1·15) | 0·97 (0·85-1·11) | 1·02 (0·89-1·17) | 1·08 (0·92-1·27) | 0·93 (0·79-1·11) | 0·81 (0·66-1·00) |
| Cancer | 0·99 (0·88-1·10) | 0·88 (0·73-1·05) | 1·01 (0·82-1·25) | 0·97 (0·77-1·24) | 1·00 (0·75-1·33) | 0·94 (0·71-1·24) | 0·87 (0·61-1·24) |
| Metabolic disorders | 1·04 (0·98-1·10) | **1·09** (1·00-1·19) | 1·15 (1·03-1·28) | 1·09 (0·97-1·23) | 1·04 (0·90-1·19) | 1·07 (0·94-1·23) | 1·03 (0·87-1·21) |

**Table S3. Mutually adjusted prevalence ratios with 95% confidence intervals for new symptom clusters with grade of impairment moderate to strong (…continued)**

| | Musculoskeletal pain | Upper respiratory symptoms | Rash/paresthesia | Hair loss | Abdominal symptoms | Nausea/vomiting | Chills/fever |
|---|---|---|---|---|---|---|---|
| Age, per 10 years | **1·30** (1·23-1·38) | **1·13** (1·04-1·22) | **1·12** (1·04-1·22) | 0·98 (0·89-1·07) | 1·05 (0·93-1·18) | 0·99 (0·85-1·16) | 1·07 (0·88-1·31) |
| Sex | | | | | | | |
| Male (ref.) | | | | | | | |
| Female | **1·55** (1·34-1·78) | **1·20** (1·00-1·45) | **1·70** (1·38-2·10) | **8·32** (5·61-12·3) | **1·54** (1·14-2·08) | **2·46** (1·55-3·89) | 1·67 (0·97-2·85) |
| University entrance qualification | | | | | | | |
| Yes (ref.) | | | | | | | |
| No | **1·21** (1·06-1·39) | **1·26** (1·04-1·52) | 1·04 (0·85-1·26) | **1·33** (1·06-1·68) | 1·15 (0·87-1·52) | 1·13 (0·77-1·66) | 1·14 (0·69-1·91) |
| Smoking status, N (%) | | | | | | | |
| Never (ref.) | | | | | | | |
| Former | 1·13 (0·98-1·30) | 1·07 (0·88-1·30) | **1·27** (1·03-1·57) | 0·98 (0·76-1·26) | 1·07 (0·79-1·46) | **1·52** (1·02-2·27) | 1·25 (0·75-2·10) |
| Current | **1·44** (1·18-1·75) | **1·57** (1·21-2·04) | **1·69** (1·28-2·24) | 1·24 (0·89-1·74) | **1·53** (1·03-2·27) | **2·14** (1·32-3·48) | **1·95** (1·02-3·73) |
| BMI, per 5 kg/m² | **1·11** (1·05-1·17) | **1·11** (1·02-1·20) | 1·08 (0·99-1·17) | 1·04 (0·95-1·13) | 0·95 (0·81-1·11) | 0·80 (0·64-1·00) | 0·90 (0·70-1·17) |
| Time since positive PCR, per month | 1·00 (0·96-1·04) | 0·95 (0·90-1·01) | 0·96 (0·91-1·02) | 0·87 (0·81-0·92) | 1·01 (0·93-1·09) | 0·92 (0·83-1·02) | 1·03 (0·89-1·19) |
| Treatment of acute SARS-CoV-2 infection | | | | | | | |
| No medical care (ref.) | | | | | | | |
| Outpatient care | **2·61** (2·27-2·99) | **2·70** (2·23-3·27) | **2·55** (2·07-3·14) | **2·44** (1·93-3·09) | **2·98** (2·22-3·99) | **2·65** (1·79-3·92) | **3·64** (2·21-5·98) |
| Inpatient care | **2·58** (2·03-3·30) | **2·55** (1·81-3·58) | **4·59** (3·38-6·23) | **5·60** (4·01-7·82) | **3·90** (2·34-6·50) | 2·21 (0·95-5·18) | **5·85** (2·75-12·4) |
| Pre-existing conditions | | | | | | | |
| Musculoskeletal disorders | **1·21** (1·06-1·39) | **1·33** (1·10-1·61) | 1·10 (0·89-1·35) | 1·08 (0·84-1·37) | 1·14 (0·86-1·53) | 1·40 (0·95-2·05) | 1·28 (0·80-2·07) |
| Cardiovascular disorders | **1·18** (1·01-1·37) | 1·14 (0·92-1·42) | **1·32** (1·04-1·67) | 1·32 (1·00-1·75) | 1·24 (0·88-1·74) | 1·41 (0·86-2·32) | 1·13 (0·66-1·93) |
| Respiratory diseases | 1·06 (0·90-1·26) | **1·60** (1·29-1·98) | **1·50** (1·20-1·88) | 1·02 (0·75-1·37) | 0·97 (0·66-1·42) | 1·05 (0·65-1·70) | 0·98 (0·52-1·86) |
| Mental disorders | **1·29** (1·10-1·51) | 1·14 (0·91-1·43) | 0·94 (0·73-1·21) | 1·02 (0·77-1·36) | **1·60** (1·14-2·24) | **1·89** (1·24-2·89) | 1·07 (0·57-2·00) |
| Neurological or sensory disorders | 1·12 (0·96-1·30) | 1·11 (0·90-1·37) | 1·21 (0·97-1·53) | 0·98 (0·75-1·30) | 0·93 (0·66-1·31) | 1·30 (0·85-1·99) | 0·77 (0·42-1·42) |
| Dermatological diseases | 0·91 (0·75-1·11) | 0·93 (0·72-1·22) | 1·07 (0·81-1·41) | 1·20 (0·88-1·62) | 0·59 (0·36-0·97) | 0·44 (0·21-0·92) | 0·69 (0·32-1·48) |
| Cancer | 0·99 (0·74-1·34) | 1·24 (0·85-1·82) | 0·82 (0·50-1·33) | 0·82 (0·47-1·43) | 1·50 (0·85-2·64) | 1·47 (0·69-3·12) | 1·49 (0·60-3·69) |
| Metabolic disorders | 1·03 (0·88-1·20) | 0·98 (0·79-1·22) | 1·10 (0·87-1·38) | **1·32** (1·03-1·70) | 1·16 (0·82-1·63) | 0·99 (0·62-1·59) | 1·25 (0·73-2·14) |

**Left panel (general health)**

| | 18-<30 years | 30-<40 years | 40-<50 years | 50-<60 years | 60-65 years |
|---|---|---|---|---|---|
| Average total loss - men | 6.7 | 7.9 | 10.7 | 11.8 | 13.6 |
| Average total loss - women | 9.4 | 10.5 | 13.4 | 14.3 | 14.8 |
| Unattributed - men | 1.27 | 1.55 | 2.06 | 2.69 | 3.10 |
| Unattributed - women | 1.69 | 1.43 | 1.54 | 2.68 | 2.53 |
| Fatigue - men | 1.51 | 1.78 | 2.84 | 2.65 | 2.99 |
| Fatigue - women | 1.58 | 2.58 | 2.80 | 1.93 | 2.19 |
| Neurocognitive impairment - men | 0.79 | 0.60 | 1.54 | 1.36 | 1.58 |
| Neurocognitive impairment - women | 0.83 | 0.90 | 1.91 | 1.91 | 2.98 |
| Chest symptoms - men | 0.88 | 0.83 | 0.91 | 1.73 | 2.71 |
| Chest symptoms - women | 1.80 | 1.44 | 1.79 | 2.23 | 2.00 |
| Smell or taste disorder - men | 1.01 | 1.06 | 0.92 | 0.52 | 0.63 |
| Smell or taste disorder - women | 1.18 | 1.26 | 1.23 | 1.29 | 0.55 |
| Anxiety/Depression - men | 0.01 | 0.54 | 0.51 | 0.79 | 0.77 |
| Anxiety/Depression - women | 0.68 | 0.25 | 1.05 | 0.78 | 0.94 |
| Headache/dizzyness - men | 0.51 | 0.46 | 0.59 | 0.66 | 0.41 |
| Headache/dizzyness - women | 0.53 | 0.54 | 0.51 | 0.66 | 0.91 |
| Musculoskeletal pain - men | 0.05 | 0.21 | -0.11 | 0.53 | 0.40 |
| Musculoskeletal pain - women | 0.37 | 0.67 | 0.80 | 0.80 | 0.53 |
| Upper respiratory symptoms - men | 0.19 | 0.29 | 0.26 | 0.33 | -0.02 |
| Upper respiratory symptoms - women | 0.07 | 0.14 | 0.14 | 0.74 | 0.57 |
| Rash/paresthesia - men | 0.13 | 0.24 | 0.47 | 0.04 | 0.32 |
| Rash/paresthesia - women | 0.07 | 0.31 | 0.63 | 0.22 | 0.47 |
| Hair loss - men | 0.01 | -0.02 | -0.01 | -0.00 | 0.17 |
| Hair loss - women | -0.03 | 0.48 | 0.08 | 0.13 | 0.37 |
| Abdominal symptoms - men | -0.04 | 0.11 | 0.15 | 0.20 | -0.04 |
| Abdominal symptoms - women | 0.02 | 0.09 | 0.04 | 0.09 | -0.09 |
| Nausea/vomiting - men | 0.22 | 0.03 | 0.08 | 0.06 | 0.34 |
| Nausea/vomiting - women | 0.21 | 0.07 | 0.32 | 0.24 | 0.09 |
| Chills/feverr - men | -0.01 | 0.10 | 0.15 | -0.10 | -0.09 |
| Chills/feverr - women | 0.17 | 0.14 | 0.17 | 0.21 | 0.24 |

**Right panel (working capacity)**

| | 18-<30 years | 30-<40 years | 40-<50 years | 50-<60 years | 60-65 years |
|---|---|---|---|---|---|
| Average total loss - men | 5.5 | 6.5 | 9.2 | 11.6 | 14.9 |
| Average total loss - women | 7.5 | 9.0 | 12.5 | 14.0 | 15.3 |
| Unattributed - men | 1.02 | 0.61 | 1.25 | 2.47 | 4.13 |
| Unattributed - women | 0.91 | 0.68 | 0.51 | 2.05 | 2.57 |
| Fatigue - men | 1.09 | 1.92 | 3.19 | 2.98 | 3.23 |
| Fatigue - women | 1.52 | 2.55 | 2.70 | 1.92 | 2.41 |
| Neurocognitive impairment - men | 1.16 | 0.88 | 1.51 | 1.56 | 1.73 |
| Neurocognitive impairment - women | 1.42 | 1.67 | 2.78 | 2.39 | 2.69 |
| Chest symptoms - men | 0.37 | 0.62 | 0.21 | 1.20 | 2.12 |
| Chest symptoms - women | 0.52 | 0.68 | 1.17 | 1.37 | 1.75 |
| Smell or taste disorder - men | 0.23 | 0.38 | 0.29 | 0.15 | 0.42 |
| Smell or taste disorder - women | 0.04 | 0.39 | 0.47 | 0.97 | 0.24 |
| Anxiety/Depression - men | 0.16 | 0.45 | 0.83 | 0.83 | 0.45 |
| Anxiety/Depression - women | 1.36 | 0.53 | 1.06 | 0.63 | 1.33 |
| Headache/dizzyness - men | 0.73 | 0.97 | 0.53 | 0.46 | 0.61 |
| Headache/dizzyness - women | 0.41 | 0.59 | 0.61 | 0.83 | 0.82 |
| Musculoskeletal pain - men | 0.07 | 0.06 | -0.02 | 0.90 | 0.79 |
| Musculoskeletal pain - women | 0.37 | 0.45 | 1.14 | 0.72 | 0.47 |
| Upper respiratory symptoms - men | 0.14 | -0.15 | 0.29 | 0.21 | -0.21 |
| Upper respiratory symptoms - women | 0.15 | 0.17 | 0.14 | 1.00 | 0.38 |
| Rash/paresthesia - men | 0.09 | 0.33 | 0.43 | 0.11 | 0.89 |
| Rash/paresthesia - women | 0.14 | 0.38 | 1.02 | 0.32 | 0.64 |
| Hair loss - men | 0.02 | -0.05 | -0.03 | -0.00 | 0.07 |
| Hair loss - women | -0.01 | 0.57 | 0.19 | 0.42 | 0.53 |
| Abdominal symptoms - men | 0.01 | 0.03 | 0.13 | 0.14 | -0.28 |
| Abdominal symptoms - women | 0.21 | 0.07 | 0.12 | 0.30 | 0.01 |
| Nausea/vomiting - men | 0.24 | 0.09 | 0.06 | 0.25 | 0.45 |
| Nausea/vomiting - women | 0.21 | 0.13 | 0.17 | 0.28 | 0.15 |
| Chills/feverr - men | 0.05 | 0.14 | 0.14 | -0.10 | 0.07 |
| Chills/feverr - women | 0.14 | 0.04 | 0.23 | 0.40 | 0.35 |

**Figure S6. Attributable percentage loss of general health (left panel) and working capacity (right panel), stratified by age categories and sex.**

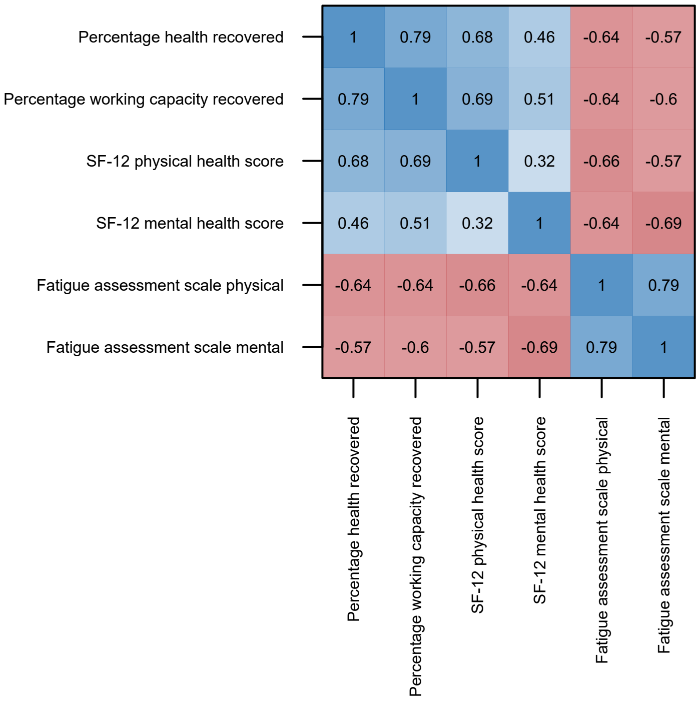

**Figure S7. Pairwise Pearson`s correlation coefficient between measures of recovered general health, working capacity, physical and mental health score (SF-12), and fatigue assessment.**

**Table S8. Participation according to age and sex.**

| Sex | Source population* | Participants (%) |
|---|---|---|
| Male | 24 959 | 4829 (19·3) |
| Female | 25 483 | 6881 (27·0) |
| Total | 50 442 | 11 710 (23·2) |

| Age class (years) | Source population | Participants (%) |
|---|---|---|
| <30 | 14 045 | 2474 (17·6) |
| 30 - <40 | 10 710 | 2158 (20·1) |
| 40 - <50 | 9504 | 2075 (21·8) |
| 50 - <60 | 11 633 | 3443 (29·6) |
| ≥ 60 | 4565 | 1560 (34·2) |
| Total | 50 457 | 11 710 (23·2) |

* n=15 unknown or non-binary sex
